# Increased entrainment and decreased excitability predict efficacious treatment of closed-loop phase-locked rTMS for treatment-resistant depression

**DOI:** 10.1101/2023.10.09.23296751

**Authors:** Xiaoxiao Sun, Jayce Doose, Josef Faller, James R. McIntosh, Golbarg T. Saber, Sarah Huffman, Spiro P. Pantazatos, Han Yuan, Robin I. Goldman, Truman R. Brown, Mark S. George, Paul Sajda

## Abstract

Transcranial magnetic stimulation (TMS) is an FDA-approved therapy for major depressive disorder (MDD), specifically for patients who have treatment-resistant depression (TRD). However, TMS produces response or remission in about 50% of patients but is ineffective for the other 50%. Limits on efficacy may be due to individual patient variability, but to date, there are no good biomarkers or measures of target engagement. In addition, TMS efficacy is typically not assessed until a six-week treatment ends, precluding the evaluation of intermediate improvements during the treatment duration. Here, we report on results using a closed-loop phase-locked repetitive TMS (rTMS) treatment that synchronizes the delivery of rTMS based on the timing of the pulses relative to a patient’s individual electroencephalographic (EEG) prefrontal alpha oscillation informed by functional magnetic resonance imaging (fMRI). We find that, in responders, synchronized delivery of rTMS produces two systematic changes in brain dynamics. The first change is a decrease in global cortical excitability, and the second is an increase in the phase entrainment of cortical dynamics. These two effects predict clinical outcomes in the synchronized treatment group but not in an active-treatment unsynchronized control group. The systematic decrease in excitability and increase in entrainment correlated with treatment efficacy at the endpoint and intermediate weeks during the synchronized treatment. Specifically, we show that weekly tracking of these biomarkers allows for efficacy prediction and potential of dynamic adjustments through a treatment course, improving the overall response rates.

## 1 Introduction

Major depressive disorder (MDD) is a debilitating and often persistent condition affecting millions of people worldwide [1, 2]. Despite receiving multiple rounds of first-line monotherapy at adequate dosage, duration, and patient compliance, up to 20-30% of individuals with MDD fail to achieve remission, leading to a diagnosis of treatment-resistant depression (TRD) [3, 4]. In recent years, research on treating TRD has gained significant attention [5– 7]. Repetitive transcranial magnetic stimulation (rTMS) is a relatively recent procedure for noninvasively stimulating the brain, particularly the prefrontal cortex. rTMS is an effective treatment for TRD with minimal side effects [8, 9]. Although considerable clinical data support its efficacy for a subgroup of TRD patients, the mechanisms of action of rTMS and the combination of its stimulation parameters, particularly individualized for each patient, remain unclear. Understanding these mechanisms and developing methods to engage them at the individual patient level will likely increase the efficacy of rTMS across the TRD population [10–12].

Recent research suggests that the delivery of rTMS causes long-term inhibition and excitation of neurons in multiple brain areas. The mechanism of these effects is believed to involve changes in synaptic efficacy akin to long-term potentiation or long-term depression [9]. As such, interest has steadily grown in using neurophysiologic measures of cortical excitability as a biomarker for the outcomes of stimulation-based treatment of MDD [13–15]. Additionally, alpha oscillations have been implicated in network connectivity, with the phase of alpha linked to activation and release of inhibition across and within networks [16–19]. Thus, an active area of investigation is whether the phase of the endogenous EEG alpha rhythms mediates top-down influences in the human brain, particularly in response to brain stimulation (e.g., TMS). Several groups have investigated synchronized TMS delivery to the alpha phase (or the mu/beta rhythm in the motor system) and have shown acute/transient effects suggesting that excitability is indexed by phase [20–22]. However, no group has examined the effects of individualized phase-synchronized rTMS applied at a conventional rTMS treatment time scale (i.e., 4-6 weeks) as a clinical intervention.

Under the hypothesis that the EEG alpha phase could act as a gating mechanism where different phases in the cycle are associated with states of low and high excitability across or within the network [18, 23], our group developed a novel closed-loop EEG-synchronized rTMS system to individualize the timing of rTMS pulse train delivery for TRD patients receiving treatment [24]. Specifically, our system synchronizes the delivery of the rTMS with ongoing prefrontal brain activity by triggering an rTMS pulse train at a specific phase of a patient’s prefrontal EEG alpha (6-13 Hz) rhythm determined by a simultaneous fMRI-EEG-TMS (fET) scan to optimally engage the therapeutic target region [23, 25, 26], which forms a strategic stimulation encompassing both spatial (targeting specific locations) and temporal (concerning other cerebral events) precision (see Fig. 1c). We conducted a randomized, active-comparator, and double-blind clinical study (ClinicalTrials.gov ID: NCT03421808 [27]) that resulted in 24 TRD patients enrolled and completing a 6-week rTMS treatment with their clinical progress evaluated weekly (see Fig. 1a). Twelve patients received closed-loop EEG-synchronized rTMS, where pulse trains were synchronized to the patient’s EEG prefrontal alpha phase (SYNC group), while the other 12 patients were treated with the same closed-loop EEG-rTMS system but with rTMS delivered at randomized phases (UNSYNC group).

**Fig. 1.**
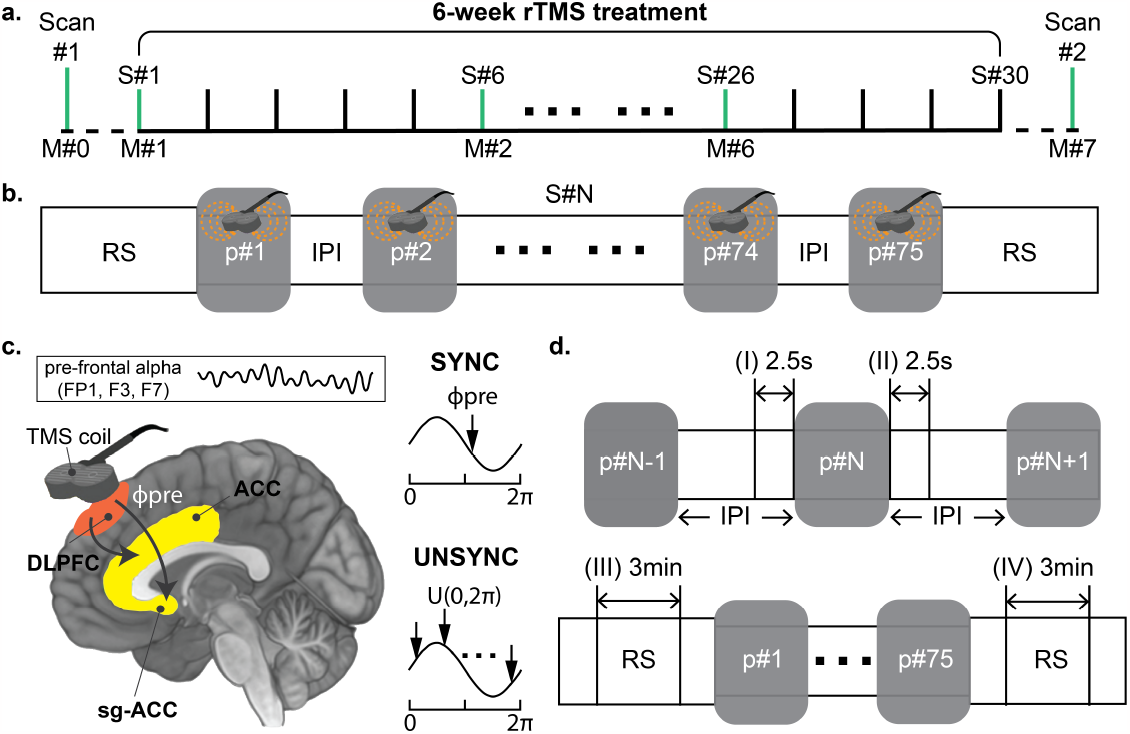
Longitudinal treatment design and time windows used for analysis. **a**. The pretreatment scan (scan #1) was done with an fMRI-EEG-TMS (fET) system to determine the subject specific “preferred” phase *ϕ*pre for each patient [23, 25, 26]. *ϕ* _pre_ is defined as the alpha phase that evoked the strongest activity in the dorsal anterior cingulate cortex (dACC) after the TMS administration [19, 26]. If the patient was assigned to the SYNC group, this phase was the target phase *ϕ*_tar_ during the treatment. Patients then received 30 rTMS treatment sessions, with the treatment efficacy clinically evaluated using the Hamilton Depression Rating Scale (HDRS). It was measured before the treatment sessions of each week (sessions with HDRS measurement are highlighted in green). After all treatment sessions, another fET scan (scan #2) was done to obtain each patient’s post-treatment preferred phase *ϕ* _post_ and a final HDRS assessment was performed. Across the entire clinical trial, there were eight HDRS measurements (M#0 to M#7); **b**. In each session, there were two 5-minute rest periods (RS) before the first and after the last rTMS pulse train. Each treatment consisted of 75 rTMS pulse trains (marked as a gray box), and in each rTMS pulse train, 40 TMS pulses were delivered at each patient’s individual quasi-alpha frequency (IAF, 6-13Hz). The IAF was estimated at the resting state before each session’s first rTMS pulse train. The interval between each rTMS pulse train is referred to as the inter-pulse train interval (IPI). **c**. The rTMS was applied over the left dorsal lateral prefrontal cortex (DLPFC, over electrode F3). Within the SYNC group, the onset of the rTMS pulse train was synchronized to the patient’s *ϕ* _pre_ (*ϕ* _tar_ = *ϕ*_pre_) to maximize neural engagement in the anterior cingulate cortex (ACC) and subgenual-ACC (sg-ACC) [18, 19, 23, 31], which is believed to be an antidepressant mechanism of TMS [12, 32, 33]. Conversely, for the UNSYNC subjects, each pulse train onset was targeted to a randomly generated phase (*ϕ*_tar_ ∼ *U* ([0, 2*π*]), each arrow indicates the target phase of one pulse train). Pre-frontal quasi-alpha oscillation was measured with EEG signal from electrodes FP1, F3, and F7 (i.e., near the rTMS target region; see SI for more details about the definition of region of interest (ROI)). These are the same channels used to determine the IAF during the first 5-minute resting period. **d**. Four time windows were used from the EEG recordings during the treatment session for analysis. For the IPI period, the 2.5 seconds before and after each rTMS pulse train was analyzed. For the resting period (RS), the middle 3 minutes of the 5-minute data was used.

This trial and a partial analysis of the effects of the magnitude of entrainment have been detailed elsewhere [25, 28]. In this paper, we consider the consistency of the entrainment effects with respect to phase and how both magnitude and phase consistency interact with cortical excitability to form a set of biomarkers that directly predict treatment efficacy. This predictive power is observed in the SYNC group but not the UNSYNC group. Specifically, we identified a decrease in global cortical excitability and increased entrainment at the near target site within the SYNC group. We established their significant associations with clinical outcomes. Furthermore, in contrast to the conventional retrospective assessment of the correlation between measurements and clinical improvement at the experiment’s conclusion, this study adopted a novel approach by integrating excitability and entrainment measurements weekly. Prospective weekly tracking of these biomarkers showed that integrated biomarker measures could be used to dynamically adjust the treatment and/or determine that a patient should be re-evaluated for another therapeutic protocol.

These results showed that synchronized treatment made it possible to track and predict patients’ improvement over time using these proposed biomarker measures. This use of closed-loop EEG-synchronized rTMS represents a significant step towards personalized medicine for treating TRD [7, 29]. By synchronizing rTMS pulse train delivery with the ongoing prefrontal brain activity of each patient with biomarker tracking, we were able to optimize treatment outcomes for individual patients, resulting in a significant improvement in response rates (85.7%) compared to conventional rTMS treatment (∼40%) reported in literature [11, 12, 30].

## 2 Results

### 2.1 Global cortical excitability decreases at the resting state with synchronized stimulation

Global cortical excitability in the individual alpha frequency (IAF, i.e., quasi-alpha band 6-13Hz, see details about IAF in the **S.2** section of the supplementary information (SI)) was measured by Global Mean Field Power (GMFP). GMFP changes (∆GFMP) between resting states (before vs. after rTMS treatment session) were compared to evaluate the cortical excitability changes for each treatment session (see time window (III) and (IV) in Fig. 1d). We observed a significant decrease in GMFP within the SYNC group (∆GFMP(SYNC)*<* 0: *p <* 0.001) where the decrease in the SYNC group was also significantly greater compared to the UNSYNC group (∆GFMP(SYNC)*<*∆GFMP(UNSYNC): *p* = 0.007, see Fig. 2a.1). When evaluated by week, we found a significant decrease in GMFP by treatment session within the SYNC group at week#1, #2, #4, and #6 (significance level was represented as the black asterisk in Fig. 2a.2, more details can be found in **Table S.4** of SI). However, no such session-level effect was observed for the UNSYNC group. This is despite the decreasing trend in global cortical excitability across treatments in the UNSYNC group (i.e., GMFP measured at week#6 after the treatment is significantly smaller than GMFP measured at week#1 before any treatment, see Fig. 2a.2).

**Fig. 2.**
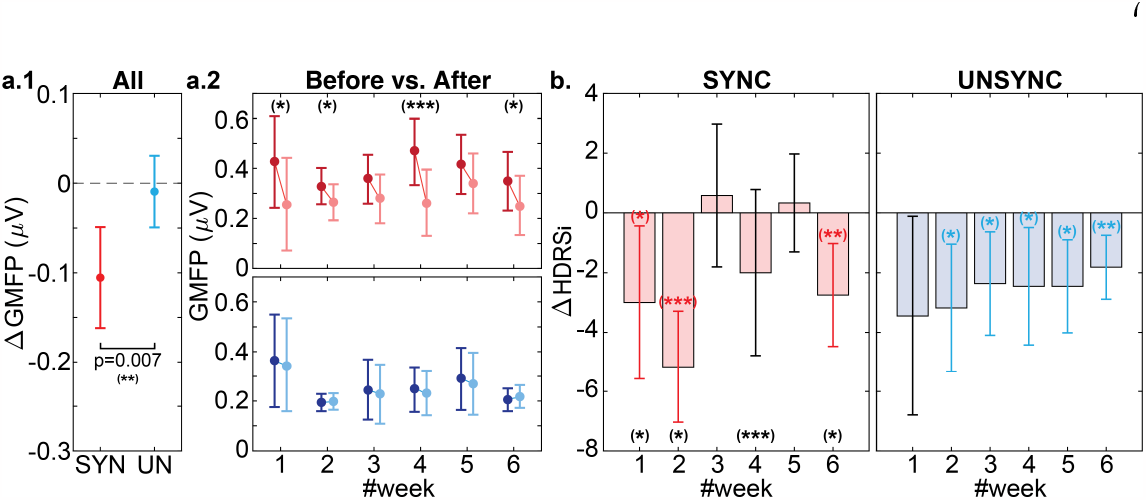
Relationship between GMFP changes and treatment outcomes. **a.1** Overall, SYNC subjects had a significantly greater GMFP decrease than UNSYNC subjects (t-test: *p* = 0.007). **a.2** In the weekly comparison, within the SYNC group (represented by red), significant GMFP decreases before (dark red) and after (light red) each treatment session were observed in weeks#1, #2 #4, and #6. (A paired t-test was used to compare the changes between ‘before’ and ‘after’ within each treatment for each week. The detailed statistical results are available in the **Table S.4** of SI. The significance level of the result is indicated by a black asterisk, where (∗ ∗ ∗) indicates *p <* 0.001, (∗ ∗) indicates *p <* 0.01, and (∗) indicates *p <* 0.05.) No significant GMFP changes by session were observed for the UNSYNC group (represented by blue). **b**. For the SYNC group, weeks with greater clinical improvement (i.e., statistically significant weekly HDRS score decreases: ∆HDRS_*i*_ *<* 0) were the weeks having significant decreases in GMFP after treatment (i.e., week#1, #2, #6. Similarly, the significance level is indicated by a red/blue asterisk, and the week with significant GMFP decreases before and after each treatment session is highlighted by the black asterisks as **a.2**). No such association between ∆GMFP and ∆HDRS_*i*_ was directly observed for the UNSYNC group in this weekly alignment.

To better understand excitability changes (i.e., ∆GMFP) between the SYNC and UNSYNC groups, we considered these changes within the context of the patients’ clinical improvement. The decrease in GMFP within the SYNC group was found to align with changes in their weekly Hamilton Depression Rating Scale (HDRS) (∆HDRS_*i*_, where *i* indicates the treatment week, see the details of HDRS in Section 4.2). Patients in the SYNC group had a significant clinical improvement (i.e., ∆HDRS_*i*_ *<* 0 with 95% confidence level) at weeks #1, #2, and #6, which occurred only when there was a significant GMFP decrease after treatment (see Fig. 2b). To further investigate the correlation between GFMP decrease and clinical response, the individual level correlation between change in HDRS and GMFP across subjects is reported in the **S.5B** section of SI. We found that GMFP change (negative value for a decrease) is significantly positively correlated with HDRS on a group-level, which means the greater the decrease in GMFP, the lower the HDRS (*p* = 0.019, see **Table S.5** in SI).

### 2.2 Stronger post-stimulation entrainment develops with synchronized stimulation

We evaluated the post-stimulation phase distribution by computing the weighted inter-trial phase coherence (wITPC). wITPC is derived from an ensemble of phase values at a particular time point in trials [25, 34]. It is a vector value in polar coordinates, characterized by its scalar magnitude ranging from [0, 1] and a specific direction within the polar angle interval of 0° to 360°. Therefore, our measurement consists of two components: the magnitude (i.e., wITPC^[1]^ ∈ [0, 1]), which measures how well the first post-stimulation wITPC peaks are aligned within each session, and the entrainment phase/angle (i.e., *ϕ*_ent_ ∈ [0°, 360°]) indicating where they align (see Fig. 3a). In previous work, where 15 subjects (7 SYNC and 8 UNSYNC) were analyzed [25], we reported that SYNC patients showed the strongest increasing wITPC^[1]^ over the near rTMS target site (electrodes FP1, F3, and F7) across sessions. This finding is reproduced when we repeat the same analysis with nine additional subjects (5 SYNC and 4 UNSYNC). To align the wITPC^[1]^ changes with weekly HDRS measurements, we plotted the changes of wITPC^[1]^ by week. We observed that this effect over the stimulation site builds over weeks, suggesting neuroplastic changes, while we saw no such effect for the UNSYNC group (see Fig. 3b1 and the **S.6C** section of SI). We also compared wITPC^[1]^ between R and NR within each group, but we did not find any significant difference between them within any group.

**Fig. 3.**
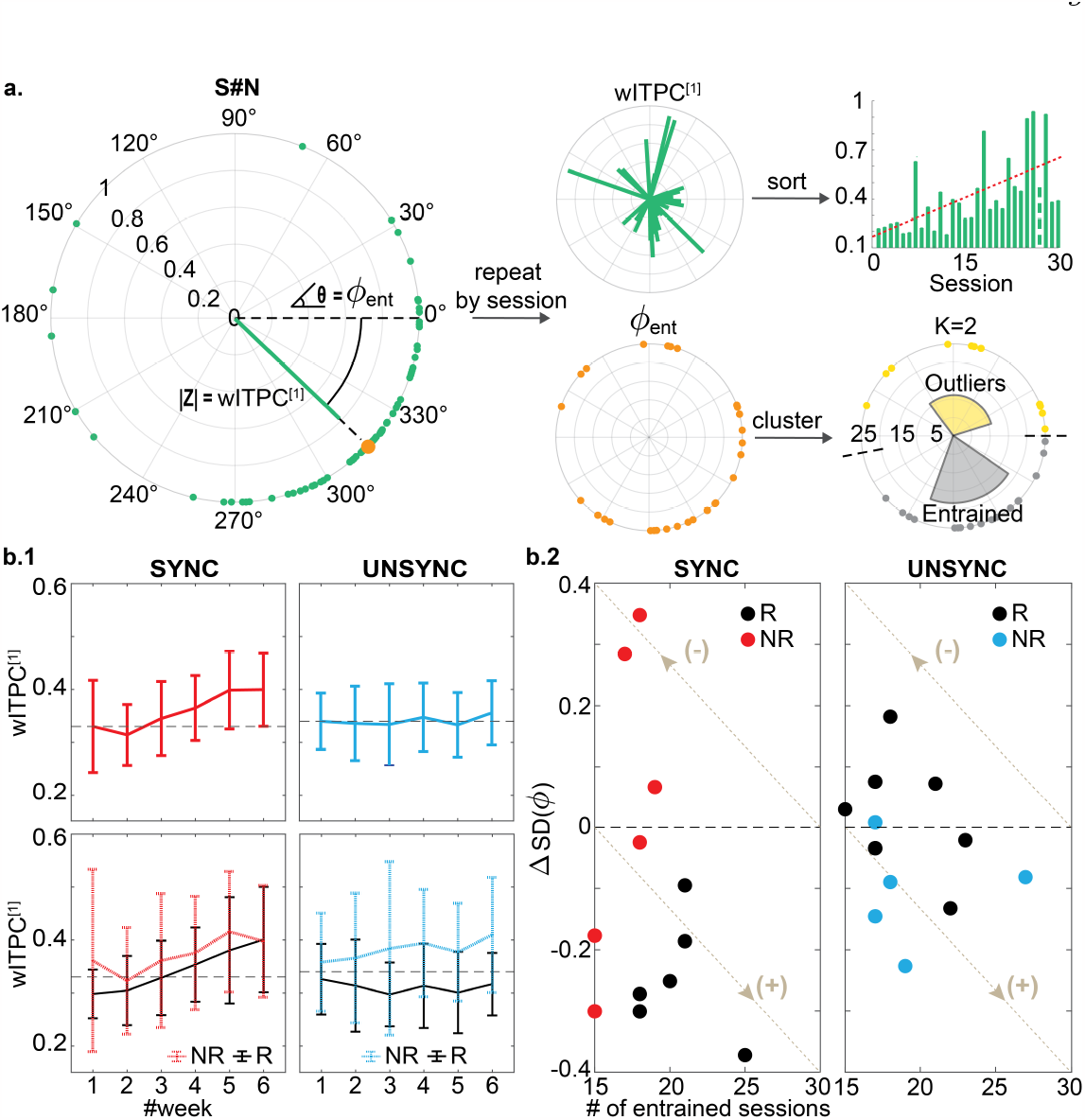
Entrainment measures within the clinical responders (R) and non-responders (NR). In the main text, we focus our analysis and report results on the near TMS target region, including electrodes FP1, F3, and F7. **a**. Flowchart of entrainment measurement calculation. Each green dot indicates one trial at the time point when the first post-stimulation wITPC peak occurs (see **S.6** section of SI for details). Two indices are computed: the magnitude wITPC^[1]^ and the phase *ϕ*_ent_. By repeating this calculation for each session for each subject, we can track two measures: (1) how the first post-stimulation wITPC^[1]^ peak changes across treatments, and (2) how *ϕ*_ent_ clusters across treatment sessions. **b.1** To align the results with the weekly HDRS assessment and reduce noise from daily variation in the measurements, we plot the weekly average of wITPC^[1]^ for each group. Within the SYNC group, there was an increase in wITPC^[1]^ across treatments, while this was not observed for the UNSYNC group (top). In addition, no significant difference between R and NR was found within each group (bottom). **b.2** Clustering results for *ϕ*_ent_. Nine out of 12 SYNC subjects and 7 out of 12 UNSYNC subjects had their entrained class with a smaller circular standard deviation than their outlier class (i.e., the difference of SD between two classes: ∆SD(*ϕ*) *<* 0), but only all clinical responders within the SYNC group have greater entrainment (3% chance this distribution would be observed, see the **S.8** section of SI for more details). Note that the more sessions having a more precise (narrower standard deviation), the better the entrainment. This direction, indicating better entrainment, follows the arrow with the **(+)** sign. Within the SYNC group, the subjects having better entrainment are also the subjects who end up being clinical responders (R, shown as black dots), while for the UNSYNC group, there is no such association between entrainment and treatment efficacy.

We then used a *K*-means-based circular clustering algorithm to characterize the consistency of *ϕ*_ent_ across treatment sessions for each patient as a function of being in the SYNC or UNYNC group. Using the Fast Optimal Circular Clustering (FOCC) method [35], we identified two classes per subject with *K* = 2. The entrained cluster/class was defined as the cluster with the most sessions, with the other being defined as the outlier cluster/class. If the number of sessions in each cluster was found to be the same (e.g., 15 out of 30 sessions in each cluster), then the cluster/class with a smaller circular standard deviation was defined as the entrained class, and the other the outlier class (see Fig. 3a) since this would indicate a more precise entrainment direction.

In Fig. 3b.2, we see that within the SYNC group, 9 out of 12 patients have an entrained class, which has a smaller standard deviation than the corresponding outlier class, compared to 7 out of 12 patients within the UNSYNC group. Although more patients within the SYNC group show strong entrainment based on these clustering results, this difference is not significant (Fisher exact test, *p* = 0.667). However, when comparing the level of entrainment itself (represented by the product of ∆SD(*ϕ*) and # of entrained sessions), we found a significant difference between groups (Mann-Whitney-Wilcoxon test: SYNC*<*UNSYNC, *p* = 0.044, see the **S.8** section of SI). Moreover, when we consider this observation within the context of the clinical results, we found that clinical responders within the SYNC group were the patients who had greater and more precise entrainment–i.e., more weeks in the entrainment class and with a smaller circular standard deviation (*p* = 0.008). No such association between properties of the entrained class and clinical improvement was found in the UNSYNC group (*p* = 0.068). More details can be found in the **S.7** and **S.8** section of SI.

### 2.3 Global excitability and entrainment as putative biomarkers that predict treatment outcome when given synchronized stimulation

Given our findings showing an association between excitability (i.e., GMFP), entrainment phase (i.e., *ϕ*_ent_ but not wITPC^[1]^) with overall clinical improvement (i.e., R/NR), we sought to investigate whether weekly changes in these measures would be predictive of treatment outcome. We created a 2-dimensional space that spanned entrainment and excitability measurements to track weekly changes of each patient relative to their weekly HDRS measurements.

For the excitability biomarker, we consider GMFP changes after the current treatment week since these were observed as transient changes that do not appear to translate to the beginning of the following week’s treatment (see Fig. 2a.2). For the entrainment biomarker, our observations were that entrainment builds progressively over time, and thus we constructed an index that is the cumulative weekly change in entrainment. With these two biomarkers, which we denote as an excitability index (EI1) and an entrainment index (EI2), we track their progression with respect to SYNC and UNSYNC groups as a function of clinical outcome (R/ NR) (see details on the calculation of EI1 and EI2 in the Methods section). Fig. 4a.1 shows the weekly changes of EI1 and EI2 for each patient. For the 12 SYNC patients shown on the left, clinical responders (R) tend to track to the upper left in this space, indicating that the weekly treatment increases entrainment and decreases excitability. There was no systematic change in these biomarkers across weeks for the UNSYNC patients. We then added the normalized EI1 and EI2 values (see Methods), creating a composite index to track weekly progress. For the SYNC group, responders had a greater value of this composite index value at week#6, relative to the non-responders, enabling a separation between responders and non-responders using this composite index. These systematic changes and separation of clinical outcomes were not observed in the UNSYNC group. Furthermore, when we aligned the final value of the normalized EI1 and EI2 with clinical improvement from the baseline (∆HDRS(%), see equation (2)), we found there was a significant positive correlation between this measure and ∆HDRS(%) within the SYNC group (*R*^2^ = 0.423, *p* = 0.022). This suggests that the degree of entrainment and excitability change correlates with the degree of clinical improvement.

**Fig. 4.**
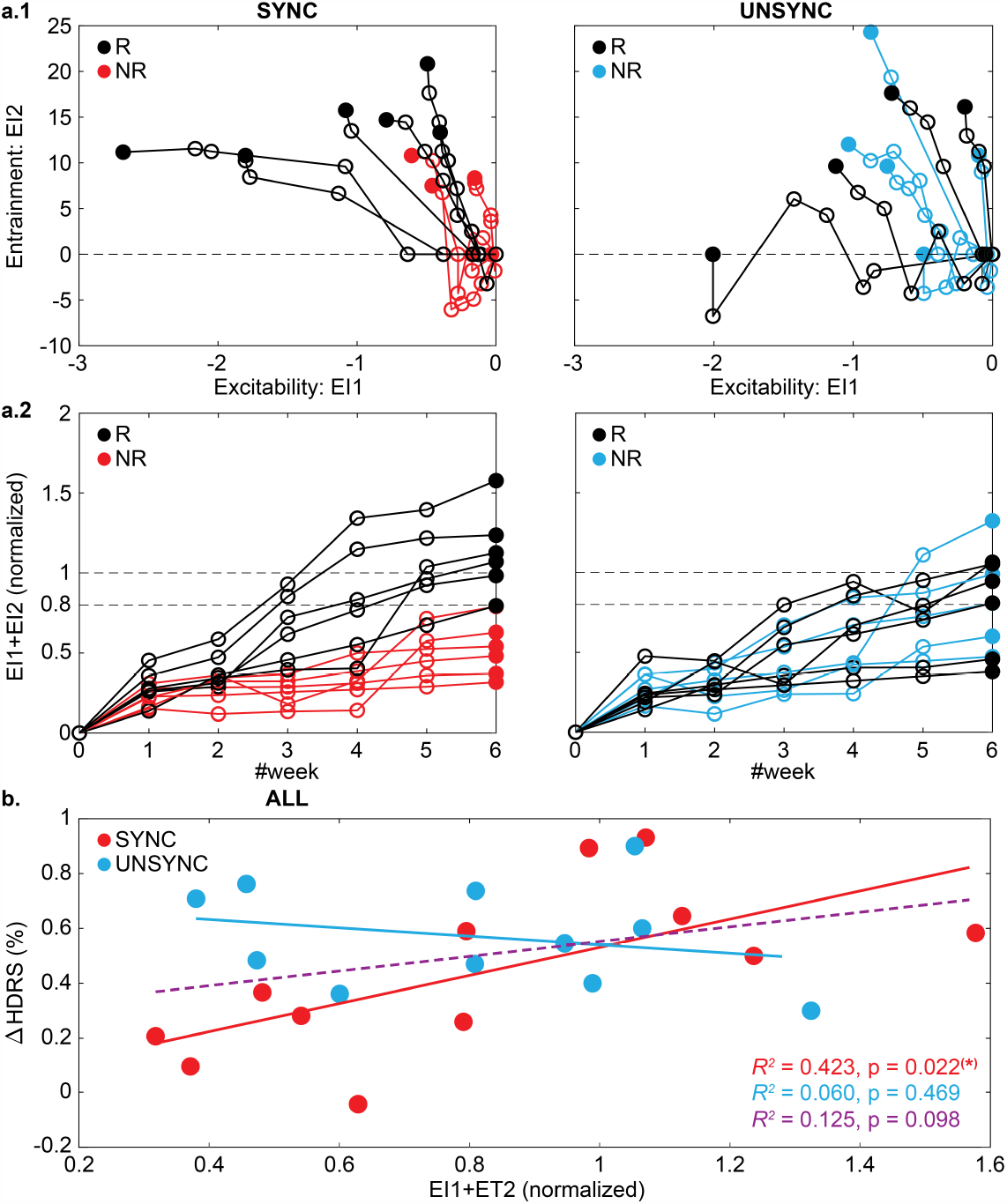
Tracking weekly changes in entrainment and excitability biomarkers. **a.1** Biomarkers, defined as an excitability index (EI1) and entrainment index (EI2), were measured each week based on the procedure described in the flowchart in Fig. 6. Each trajectory represents one patient, with each dot indicating a treatment week. The filled dot indicates the last week of the treatment (i.e., week#6). Similar to the GMFP and *ϕ*_ent_, for SYNC patients, clinical responders (R) are those that had low excitability, higher entrainment or both (i.e., smaller EI1 and/or greater EI2 at the last week). This systematic change across weeks was not observed in the UNSYNC group. **a.2** To relate EI1 and EI2 on the same scale, we take the absolute value of EI1 and normalized the two indices to [0, 1]. We then sum these normalized measures to create a single composite biomarker score. Patients with the highest composite score were ultimately clinical responders for the SYNC group. Additionally, in several cases, this separation between R and NR within the SYNC group occurs beginning in week#3. No separation between R and NR was observed in the UNSYNC group. **b**. Correlations between the composite biomarker and ∆HDRS(%) within each group (SYNC:red and UNSYNC:blue) and across all subjects (purple) were shown. There is a significant positive correlation observed within the SYNC group.

### 2.4 Specialize treatment plan based on weekly changes in global excitability and phase entrainment

Further inspection of Fig. 4a.1 and a.2 shows that changes in these biomarkers occur before the end of treatment, with a divergence between responders and non-responders being evident as early as week#3. Instead of a retrospective assessment of the correlation between measurements and clinical improvement at the study’s conclusion (especially clustering algorithm was only applied at the end of all sessions to determine which session belonged to the entrained class), we further explored the potential utility of the biomarkers. Our investigation aimed to determine whether these biomarkers could predict the likelihood of a favorable treatment response within the SYNC group or indicate the need for early re-evaluation in cases of non-response. To do this, we developed a procedure for determining a personalized treatment plan based on the individual’s progressive changes in these neural markers (see Fig. 5a).

**Fig. 5.**
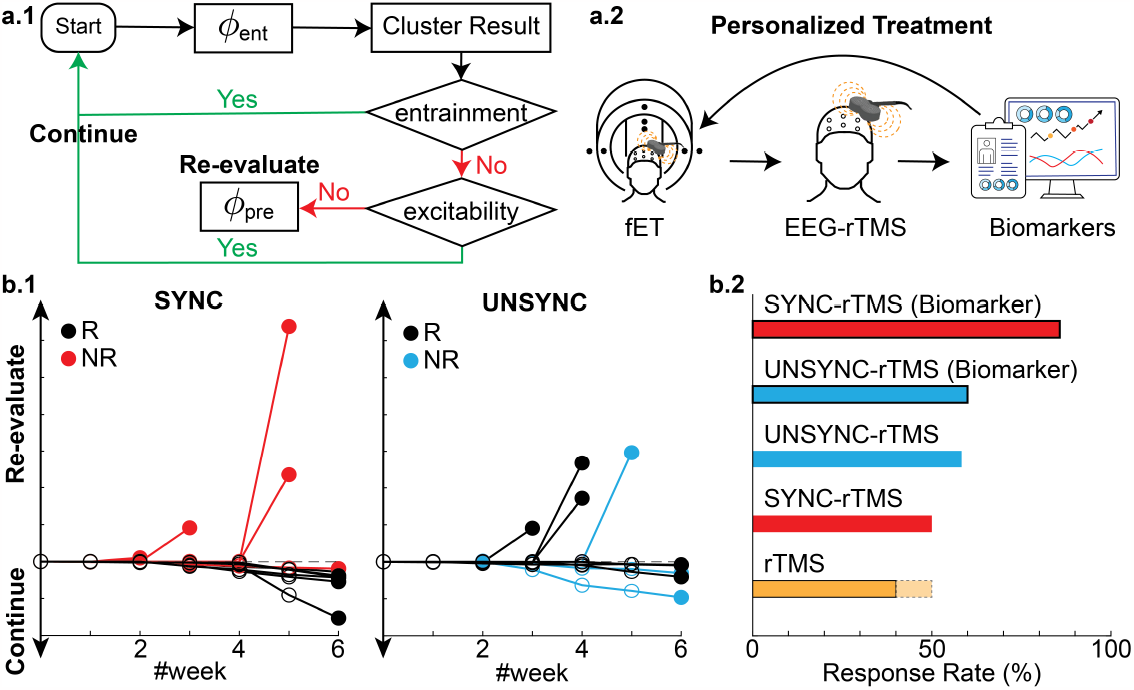
Tracking weekly biomarkers to dynamically evaluate the likely efficacy of EEG synchronized rTMS. **a.1** Decision diagram that uses entrainment and excitability biomarkers. After each week, the history of *ϕ*_ent_ of each session (no information about the future is used *ϕ*_ent_) was calculated and clustered with *K* = 2. Based on the clustering outcome, two types of effects in phase entrainment were measured (see **S.9** section in SI). First, the entrainment effect was calculated to measure how well the phases are aligned in the current entrained class. Secondly, the flipping effect is calculated, which measures the stability of this classification result. Specifically, it measures how many sessions have changed regarding the clustering result compared to the previous week (entrainment*>*filpping). If the entrainment effect is stronger than the flipping effect (meaning the patient’s sessions are consistent–i.e., similar phase direction–after the current week’s treatments), then there are no recommended changes to the treatment. When the entrainment effect is weaker than the flipping effect, the excitability biomarker is evaluated to determine if its excitability is continuing to decrease (∆EI1 ≥ 75%). If so, then then the current treatment plan is continued. For those patients who do not meet these criteria, the current treatment plan would be re-evaluated with possible actions, including 1) doing another fET session to determine if a phase shift occurred and a new phase should be dialed into the closed loop treatment or 2) stop the current rTMS treatment since the patient does not seem to be responding. **a.2** Overview of the personalized rTMS treatment. Our treatment protocol personalized the rTMS treatment concerning three key aspects: 1) Each patient underwent a simultaneous fET scan to determine their subject-specific preferred alpha phase *ϕ*pre that evoked the strongest activity in the dACC. 2) In every treatment session, we measure the IAF and deliver rTMS at the patients’ daily IAF frequency (personalized stimulation frequency, see **S.2** section for more details). Additionally, the onset of rTMS delivering is phase-locked to *ϕ*pre. 3) EEG-synchronized rTMS therapy can help to build a personalized treatment plan based on an individual’s progressive brain biomarker changes and predict a patient’s clinical outcomes before the clinical evaluation, **b.1** Results of applying this biomarker tracking and evaluation to SYNC and UNSYNC patients. The y-axis is the combined effect of phase entrainment (entrainment+flipping) and excitability. Each line represents one patient (initial session starts at 0), and each dot shows the weekly evaluation based on this biomarker tracking. **b.2** Response Rate (%) of different rTMS treatment methods. The conventional 4-6 week treatment typically only has, on average, a 40% response rate (upper bound 50%) [11, 12, 30]. Biomarker tracking can improve the response rate to 85.7% when using the EEG-synchronized rTMS treatment.

In this treatment plan, only the entrainment phase of each session until that treatment week was calculated and evaluated by the clustering algorithm (*K* = 2). Then, based on the clustering outcome, two types of effects were measured: 1) an entrainment effect, measuring how well the phase aligns with the current entrained class; 2) a flipping effect, measuring the stability of the clustering/classification result. The “flipping effect” can be viewed as measuring how many sessions caused a change in the clustering result compared to the previous week. Comparing these two effects provides a measure of the consistency of the phase synchronization given the weekly treatment. When the entrainment effect is greater than the flipping effect, the treatment is expected to progress, and clinical improvement is expected. However, if the entrainment effect is weaker than the flipping effect, the excitability should be evaluated to determine if there is a significant decrease (∆EI1_*i*_ ≥ 75%). If excitability is decreasing, the patient stays under the current treatment plan although no great entrainment effect was observed. For those patients who do not meet these criteria, the procedure would call for stopping their current treatment and re-evaluating since the biomarkers do not suggest a significant clinical improvement. Note that this procedure and all calculations are based on the current week and past measurements from the patient and do not use future information past the current week – i.e., they are truly predictions/forecasts. Applying this simple biomarker-based procedure to our data shows that five patients should be re-evaluated at some point during the 6-week treatment. Among the seven subjects recommended to continue treatments based on their weekly biomarkers, 6 are responders at the treatment endpoint. The one non-responder that would have been recommended to continue is, in fact, the only patient who did not finish all treatments in the last week (3 out of 5 treatments missed). Since entrainment is cumulative, the later sessions would more significantly affect the overall clinical outcome. In addition, the patient has the lowest combined effect (entrainment+flipping) among the seven patients. The biomarker-based causal procedure improves the response rate to 85.7% (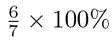, see Fig. 5b).

## 3 Discussion

The present study aimed to investigate the association between neurophysiological measures and clinical improvement in patients with MDD who underwent the personalized closed-loop phase-locked rTMS therapy. In our EEG-synchronized systems, our triggering marker in EEG is informed by an fMRI scan, where we link the EEG alpha phase with corresponding BOLD activation at the deeper brain region. It solves the technical difficulties in non-invasive neuromodulation tools such as TMS, where the ultimate therapeutic target site might be deep in the brain while the stimulation site is located superficially [23] so as to form a strategic personalized stimulation achieving both spatial and temporal precision.

First, we investigated the effects of synchronized versus unsynchronized rTMS on global cortical excitability at the resting state. Specifically, we measured GMFP in the IAF (± 0.5Hz) before and after each treatment session to evaluate cortical excitability changes. Our findings showed a significantly greater decrease in GMFP within the SYNC group than the UNSYNC group, indicating that synchronized TMS led to greater decreases in cortical excitability at the resting state after the treatment. Interestingly, this decrease in GMFP within the SYNC group aligned with patients’ clinical improvement, as measured by changes in the HDRS. This finding suggests that the decrease in global cortical excitability induced by synchronized TMS may be a potential biomarker for predicting treatment response in patients with depression. Furthermore, our results also revealed that GMFP decrease is significantly positively correlated with HDRS decrease, regardless of the treatment group. This finding also supports the conclusion that individual differences in global cortical excitability can predict treatment response in MDD patients [13]. This could have significant implications for developing personalized medicine approaches in this population. The observation of a difference in global cortical excitability in this study also highlights the potential of resting-state EEG data as a predictive tool for clinical improvement, as reported in previous investigations [15, 36].

By computing the wITPC, our study evaluated the post-stimulation phase modulation and found that the SYNC group showed stronger post-stimulation entrainment compared to the UNSYNC group. We also found that the effect of post-stimulation entrainment in magnitude (represented by wITPC^[1]^) over the stimulation site builds over weeks within the SYNC group, which suggests progressive neuroplastic changes. The distribution differences of *ϕ*_ent_ across sessions between the SYNC and UNSYNC groups were investigated by analyzing the circular clustering outputs. The SYNC group showed greater entrainment in phase based on the clustering results. Moreover, we found that clinical responders within the SYNC group were the patients who had better entrainment in *ϕ*_ent_.

Based on these findings, we developed two indices, the excitability index (EI1) and the entrainment index (EI2), to measure weekly changes in excitability and entrainment. We found that clinical responders within the SYNC group either had higher entrainment and/or lower excitability, and this difference could be observed since week#3. A biomarker representing a combined measure of normalized EI1 and EI2 showed a significant correlation with the change in HDRS, but this association was observed exclusively within the SYNC group. This finding parallels the observation that the specific modulated functional connectivity changes (e.g., between L-DLPFC and right hemisphere orbitofrontal cortex) induced by rTMS treatment are only associated with the clinical outcome in patients who received synchronized stimulation [37].

Overall, our proposed treatment protocol could be used to personalize the closed-loop EEG-rTMS treatment. Though the current results are a retrospective analysis of the decision criteria outlined in Figure 5, we are evaluating the decision criteria prospectively in a clinical trial. In this prospective clinical trial, we will track the biomarkers and reassess the preferred phase if the biomarkers do not reflect the patient’s likely response. If the preferred phase has changed, we will use this new phase to continue treatment; else, we will assign the patient to be an unlikely responder category. Our method pushes forward personalized medicine [29] and generates superior clinical outcomes than conventional rTMS treatment [11] and even invasive neurostimulation tools like Deep Brain Stimulation (40% to 70% response rate [38]). Moreover, as the field of rTMS experiences rapid expansion and integration into the therapeutic landscape of other neuropsychiatric disorders like OCD and schizophrenia [39, 40], our approach presents a compelling and personalized alternative for administering rTMS treatment to individuals afflicted.

It should be noted that the current study has several limitations that need to be addressed in future research. For example, our study was not designed to determine whether any randomly chosen phase, rather than the predetermined subject-specific preferred phase, would accomplish the same effect as long as the target phase is fixed across the treatment sessions. Future studies with adapted designs are needed to test whether rTMS can induce entrainment also if *ϕ*tar is fixed instead of tuned to each subject (see the **S.7** section in SI for more details). Moreover, the current study only included a relatively small sample of patients with depression, which may limit the generalizability of our findings to larger populations. In addition, the developed biomarkers were initially derived from the same dataset subsequently employed to demonstrate their efficacy. To address this potential source of bias, we intend to rigorously evaluate the developed biomarkers by subjecting them to validation within an independently collected dataset. Thus future studies with larger sample sizes are needed to replicate and extend our findings. Lastly, in contrast to the focus on optimizing engagement with specific brain regions, another active research area in closed-loop stimulation involves directly targeting triggers within complex disease-state-relevant brain networks [41, 42]. Therefore, further investigation of closed-loop stimulation based on phase-dependent functional connectivity changes [23, 37] represents a valuable avenue for enhancing the efficacy of MDD treatment. Nonetheless, our results provide compelling evidence that cortical excitability and entrainment biomarkers can be used to track the clinical efficacy of personalized closed-loop EEG-rTMS for treating TRD.

## 4 Methods

### 4.1 Subjects

We conducted a randomized, double-blind, active comparator-controlled clinical trial (ClinicalTrials.gov ID: NCT03421808 [27]). Clinical results were revealed only after the completion of treatment for all patients. Out of 32 enrolled patients, twenty-four patients (SYNC: 12 patients (3M, 9F), 44.0 ± 12.5 yrs; UNSYNC: 12 patients (4M, 8F), 42.3 ± 12.4 yrs) were able to complete the 6-week rTMS treatments (see the **S.1** section of SI for more details). The Institutional Review Board of Medical University South Carolina (SC, USA) reviewed and approved this study. Written informed consent was obtained from all participants before enrollment. All patients were randomly assigned to the SYNC or UNSYNC group before the rTMS treatment, and all EEG data was collected at the Medical University of South Carolina.

### 4.2 Experiment design

The longitudinal experiment design is shown in Fig. 1. Before recruitment, all patients were screened for all inclusion and exclusion criteria described in [27], [25], and [28]. Then the pre-treatment scan (scan#1) was done with our integrated fMRI-EEG-TMS (fET) system to determine the pre-treatment preferred phase *ϕ*_pre_ of each subject [19, 25]. For patients assigned to the SYNC group, the *ϕ*pre measured for each patient was used as their target phase *ϕ*tar for the entire closed-loop EEG-rTMS treatment (i.e., *ϕ*_tar_ = *ϕ*_pre_). In contrast, for the UNSYNC group, the *ϕ*_tar_ for each pulse train for a given patient was randomly drawn from a uniform distribution (i.e., *ϕ*_tar_ ∼ *U* (0, 2*π*)). This ensured an unsynchronized delivery of rTMS pulse trains unrelated to the patient’s preferred phase as assessed by the fET system. During the treatment period, each patient received a total of 30 rTMS treatment sessions (one treatment per weekday for six weeks). Within each session, there were two 5-minute resting state EEG recordings (before the first and after the last rTMS pulse train) and 75 rTMS pulse trains. In each rTMS pulse train, 40 TMS pulses were delivered at the individual’s IAF (3000 pulses/session, see details about IAF in SI), where the first pulse of each rTMS pulse train was phase-locked at *ϕ*_tar_. When all 30 treatments were completed, another scan (scan#2) was done to obtain the post-treatment preferred phase *ϕ*_post_ (Experiment summary is provided in section **S.1** to **S.3** of SI. Details about the fET system and closed-loop EEG-synchronized system are also available in [23, 25, 26]). For the evaluation of treatment efficacy, before the first treatment of each week, patients had the Hamilton Depression Score (HDRS: Ham-D 28 item -video-taped) assessments performed by a qualified individual who was blinded to the patient’s assigned treatment group (see Fig. 1a, M#N indicates one clinical assessment. There are eight assessments across the entire treatment) [28]. Equation (1) evaluates the weekly clinical improvement. A clinical responder is generally defined as a patient who has a 50% or more decrease in symptoms after the entire treatment sessions (M#7) from baseline (M#0) on the HDRS measurement (see equation (2)) [30].

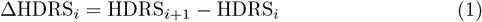

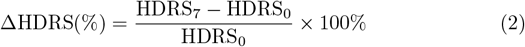

where *i* indicates the treatment week (*i* ∈ {1, 2, 3, 4, 5, 6}). HDRS_0_ represents the HDRS measured at the first fET scan (M#0) before any treatments, while HDRS_7_ is the HDRS measured at the second fET scan (M#7), which occurs at the completion of all treatment sessions.

### 4.3 EEG data analysis

#### 4.3.1 Preprocessing of EEG data

An EEG cap with 32 active sensors was used, and data were sampled at 10 kHz using a biosignal amplifier (Brain Products GmbH, Munich, Germany [43]). Before analysis, a double exponential model was fit and subtracted from the post-pulse response for all pulses in a session to suppress slow instantaneous TMS artifacts in the EEG (Note: the EEG data within the TMS pulse trains was not analyzed in our study). The entire EEG session was then low-pass filtered with a cut-off at 50 Hz and down-sampled to 250 Hz. Infomax-based Independent Component Analysis (ICA) [44] was then performed on each session for each subject independently. The CORRMAP plugin [45] was used to identify ocular artifacts across sessions, and those components were subsequently removed from the EEG data (more details about data recording and preprocessing are available in [25]). EEG data were then re-referenced to electrode location TP10 (close to the right mastoid).

Resting-state (RS) and inter-pulse train interval (IPI) data were analyzed. For each period, the EEG recording was segmented into two separate datasets for comparison purposes (RS: before and after each treatment session; IPI: before and after each rTMS pulse train, see definitions in Fig. 1d). In total, we obtained 75(trials) ×2(epochs) × 30(sessions) = 4500 epochs per patient for the IPI period and 2(epochs) ×30(sessions) = 60 epochs per patient for the RS period.

#### 4.3.2 Measurements of excitability

Since oscillatory voltage amplitude is a principal measurement that can directly reflect cortical excitability, changes in excitability can be indicated by the changes in EEG field power [14]. The cortical excitability was measured using the Global Mean Field Power (GMFP) and Local Mean Field Power (LMFP) index (see equation (3)). GMFP is often used to measure the global excitability in studies of non-invasive neuromodulation treatments (such as TMS) [13, 46]. LMFP was also measured and compared to further isolate specific contributions of different regions of interest (ROIs) to the global cortical excitability [14]. In our study, GMFP (unit: *μ*V) was computed on the TMS-evoked potentials (TEPs) of all 31 channels, and LMFP was calculated following the same procedure but from four selected ROIs (near target, contralateral to the target, media-frontal and occipital region respectively, see SI for details).

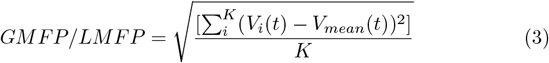

where *t* is time; *K* is the number of included channels; *V*_*i*_ is the voltage in channel *i*; *V*_*mean*_ is the mean of the voltages in all channels.

For the IPI period, both GMFP and LMFP were computed from the end of each rTMS pulse train to 2500 ms after the end of the train. TEPs were computed by averaging single trials and bandpass-filtering them with frequency band centered at IAF (IAF ± 0.5 Hz). TEPs were baseline corrected between -2500 and 0 msec before the first TMS pulse of the rTMS pulse train. For comparison purposes, 500 ms time windows were applied to the result (see **Fig. S.1**). As an index of global (local) cortical excitability, GMFP (LMFP) was computed as averaged TEPs of 31 (3) channels for ten temporal windows: -2500 to -2000 ms, -2000 to -1500 ms, -1500 to -1000 ms, –1000 to -500 ms, –500 to -128 ms, 128to 500 ms, 500 to 1000 ms, 1000 to 1500 ms, 1500 to 2000 ms and 2000 to 2500 ms, with 0 ms corresponding to the entire rTMS pulse train. The first 128ms after and the last 128ms before the rTMS pulse train were not analyzed due to filtering artifacts caused by zero padding at the beginning and end of the epoch [25]. This time window was determined by the order of the filter (order: 32) and the sampling rate (250 Hz). The results of the GMFP/LMFP calculated at the IPI period are available in the **S.5** section of SI.

Similar calculation steps were applied for the RS period, except that we applied the sliding window technique to take all 3-minute resting state data into account. A window of 2500 ms length (consistent with the time window analyzed for IPI period) was slid across the data step-wise with 1000 ms overlap, then the average of all subsets was taken as the TEPs used for the resting state. TEPs at the after-treatment resting state were baseline-corrected by the before-treatment resting state. Because the GMFP calculated at two resting states is relatively time-invariant, the average across 2500 ms is directly used to represent the global cortical excitability at the resting state of one session. The result of the GMFP measured at the RS period is presented in the main text, and the result of the LMFP measured at the RS period is available in the **S.5** section of SI.

#### 4.3.3 Measurements of entrainment

Inter-trial phase coherence (ITPC) measures the consistency of post-event oscillatory activity [47]. It reflects the temporal and spectral synchronization of EEG signal, elucidating the extent to which underlying phase-locking occurs [48]. By computing phase relationships across single trials, ITPC can be used to examine the post-stimulation cortical phase synchrony within certain areas (e.g., single electrode). ITPC values closer to 0 indicate weak phase synchrony among the trials at a particular time point *t*_*n*_, while an ITPC value closer to 1 indicates a high alignment of phase angles across trials at that point [25, 49]. Our study adjusted the calculation of ITPC with a power-based weighted average method to correct the trial weight by phase estimation accuracy. The details of the weighted ITPC (wITPC) calculation are available in the **S.6** section of SI (see equation (4), method has also been described in [25, 26]). The first post-stimulation wITPC peak (i.e., wITPC^[1]^) was defined as the first local maximum of the ITPC following the last TMS pulse in a train (see the **S.6B** section of SI).

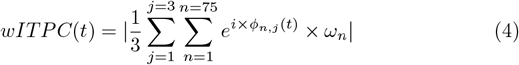

where *j* indicates the channel; *n* indicates the number of trial within one treatment session; *ϕ*_*n,j*_(*t*) is the instantaneous phase of channel *j* from trial *n* at time *t*; *ω*_*n*_ is the trial weight for trial *n* calculated based on its relative alpha power measured before stimulation (see **Fig. S.2** in SI).

We investigated the synchronization between cortical areas (e.g., between electrodes), and thus also measured synchronization via the phase-locked value (PLV). PLV estimates how the relative phase between areas is distributed over the unit circle [50]. Similar to ITPC, PLV is a scalar value that ranges from [0, 1] and is derived from an ensemble of relative phase values between electrodes across a particular time period *T*. If there is a strong phase synchronization between regions/electrodes, the relative phase occupies a small portion of the circle, and the PLV would close to 1. Conversely, regions/electrodes that are not synchronized will have their relative phase spread across the unit circle, resulting in a PLV close to 0. Equation (5) provides the formula for the PLV calculation. Because the significant differences are mostly observed within the NR group, PLV is not used as a predictive biomarker in our study to improve the response rate. The results of PLV measurements are included in the **S.10** section of SI.

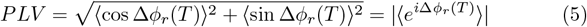

where ⟨·⟩ indicates time average; ∆*ϕ*_*r*_ is the relative phase difference between two areas (i.e., electrodes in our analysis); *T* is the time period that we are interested in (e.g., 2.5 seconds).

#### 4.3.4 Framework to measure weekly putative biomarkers of the excitability index (EI1) and entrainment index (EI2)

To evaluate the weekly changes in excitability and entrainment, we designed two indices, EI1 and EI2, to track progressive changes in GMFP and *ϕ*_ent_, respectively. Based on the result of the generalized linear mixed model (GLMM) with GMFP, we found that not only the decreases in GMFP (∆GMFP) but also the GMFP at the resting state after the treatment (GMFP To evaluate the weekly changes in excitability) correlates with the HDRS changes (see the **S.5B** section of SI for details). Thus EI1 captures the effect from both ∆GMFP and GMFP_after_ (see Fig. 6a.1). For EI2, where we track the changes of *ϕ*_ent_ via clustering since more sessions located at a narrower direction indicate better phase entrainment, both the number of sessions in the entrained class and its circular standard deviation are included in the EI2 calculation (see Fig. 6a.2). EI1 and EI2 were normalized to the scale between 0 and 1 to account for their numerical differences (e.g., Fig. 4a.2) and their values were tracked accumulatively across the weekly sessions.

**Fig. 6.**
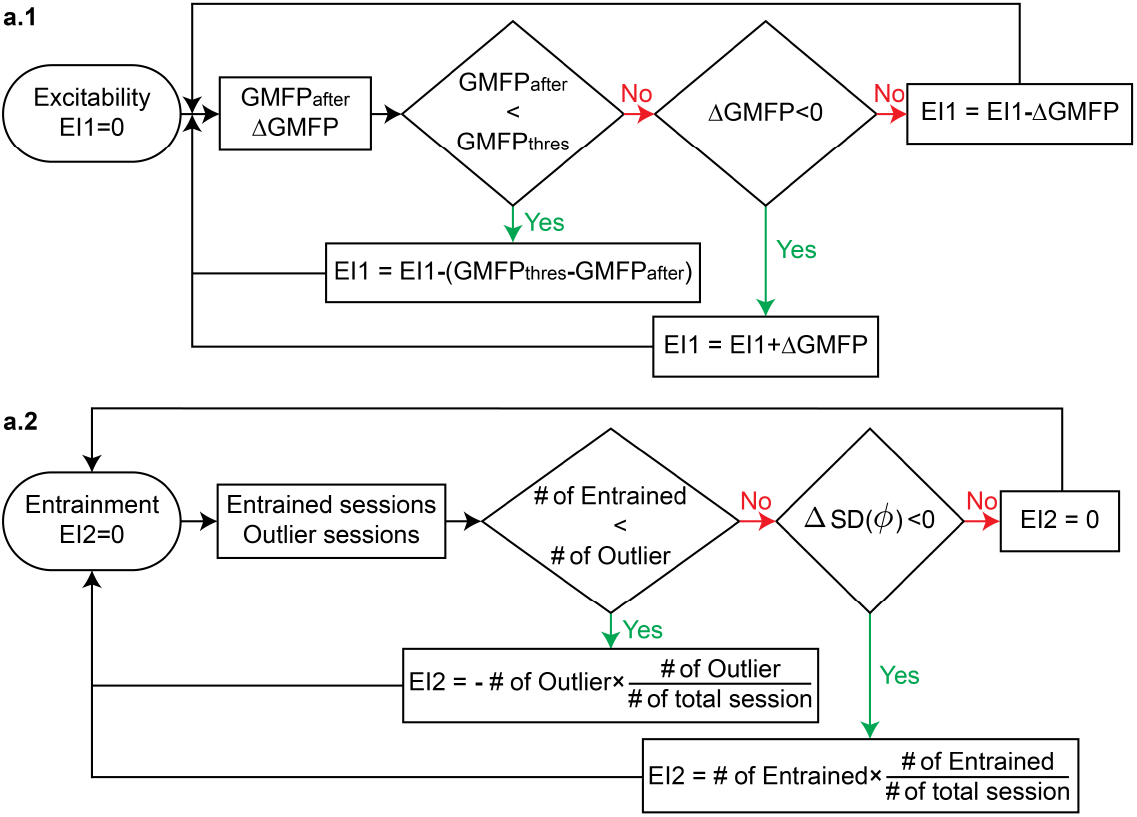
Flowchart of excitability index (EI1) and entrainment index (EI2) calculation. **a.1** EI1 was calculated based on GMFP_after_ and ∆GMFP and a smaller EI1 indicates a greater decrease in global cortical excitability. The initial EI1 value was set to 0, and after each week, both weekly averages of GMFP_after_ and ∆GMFP were calculated and used for comparison. If GMFP_after_ is smaller than the threshold value GMFP_thres_ that is computed by averaging the GMFP_after_ of all clinical responders, we interpret that the excitability remains low, and we decrease the value of EI1 by the difference between GMFP_after_ and GMFP_thres_. If not, we check for a decrease in GMFP (∆GMFP*<* 0) and update the value of EI1 based on this result. **a.2** EI2 was calculated based on the # of entrained sessions and ∆SD(*ϕ*). A greater EI2 indicates better entrainment. Before the beginning of treatment, the EI2 value was initialized to 0. For each week, we check how many sessions belong to the entrained/outlier class (note: based on the clustering result of all treatment sessions). Based on the distribution of entrained and outlier classes, we update EI2 as per the formulas (see text).

## Data Availability

All data produced in the present study are currently unavailable, but will be available upon reasonable request to the authors later.

## Acknowledgments

This work was funded by the National Institute of Mental Health (MH106775), a Vannevar Bush Faculty Fellowship from the US Department of Defense (N00014-20-1-2027), and a Center of Excellence grant from the Air Force Office of Scientific Research (FA9550-22-1-0337). We want to thank Daniel Cook and Joshua B. Teves for their help with initial data collection with closed-loop EEG-rTMS, Michael Milici for his help with building the safety circuit box and ActiChamp testing, DeeAnn Guo for her help with ActiChamp testing and initial EEG data collection, and Yida Lin for his help with initial work to integrate the closed-loop EEG-rTMS system.

## Notes

### Competing Interest Statement

Dr. Paul Sajda is a scientific advisor to Optios Inc. and OpenBCI LLC.

### Clinical Trial

NCT03421808

### Clinical Protocols

https://clinicaltrials.gov/study/NCT03421808

### Author Declarations

The Institutional Review Board of Medical University South Carolina (SC, USA) reviewed and approved this study.

