## Supplemental Information for "Increased entrainment and decreased excitability predict efficacious treatment of closed-loop phase-locked rTMS for treatment-resistant depression"

The supplementary materials accompanying this study provide additional information and details about the methods, analyses, and results presented in the main text. They include figures (Fig. **S.1** to **S.5**) and tables (Table **S.1** to **S.8**) that support and extend the findings reported in the main text. The supplementary materials also provide detailed descriptions of the procedures used in the study, including the EEG and fMRI data acquisition and analysis methods, as well as the statistical analyses performed on the data. The information contained in the supplementary materials will be of interest to researchers and clinicians interested in the use of personalized rTMS therapy for treating depression, as well as those interested in the development of EEG-synchronized stimulation protocols.

### S.1. PATIENTS

The inclusion and exclusion criteria of patient recruitment are available in [1–3]. To ensure that the baseline level of depression severity was stable at the time of study enrollment, patients were dropped from the study if they showed more than 30% improvement in the HDRS score from the time of their initial screening to the baseline assessment. The number of patients in every group, average age, gender, and average (± standard deviation) HDRS measured at pre-treatment scan (scan#1: $HDRS_{baseline}$) are shown in Table S.1. There is no significant difference between the SYNC and UNSYNC groups in age, gender, or baseline HDRS score.

**Table S.1.** Patient demographics. The number of patients in each group, age (years), sex, and $HDRS_{baseline}$ (i.e., HDRS measured at the pre-treatment scan) are shown.

| SYNC | | | | UNSYNC | | | |
|---|---|---|---|---|---|---|---|
| $N$ | Age (yrs) | Sex | $HDRS_{baseline}$ | $N$ | Age (yrs) | Sex | $HDRS_{baseline}$ |
| 12 | 44.0±12.5 | 3M, 9F | 29.7±4.3 | 12 | 42.3±12.4 | 4M, 8F | 29.9±7.0 |

### S.2. INDIVIDUAL ALPHA FREQUENCY

Individuals' electroencephalogram (EEG) can exhibit significant variability in their features, including the frequency of the alpha oscillation. Due to the non-stationary nature of EEG data, these features can vary within and between days for an individual, which poses difficulties in real-time phase tracking during treatment.

Patients' EEG quasi-alpha peaks and power stayed fairly consistent across treatment sessions. However, to account for daily variability to optimize system performance, the individual alpha frequency (IAF) and triggering threshold were recalculated at the start of every daily treatment session. We determined the IAF of each treatment session by using the 5-min resting-state recording before the treatment (eyes open), picked from a range between 6 and 13 Hz based on baseline EEG power (see [2, 4] for more details). This recording was further used to optimize individual thresholds for triggering TMS, represented by a model fit parameter, Root Mean Square Error (RMSE). This optimization was done to ensure that the system did not take longer than approximately 5 seconds to identify an appropriate EEG target phase, which ensures that delivering 3000 TMS pulses in one treatment session would take at most approximately 30 minutes. The IAF and RMSE of each patient across treatment are given in Table S.2.

### S.3. EEG-RTMS TREATMENT SESSIONS

In our study, every treatment session started and ended with a 5-minute resting-state recording of EEG (except for one UNSYNC patient, sub ID:sub-16, whose resting-state data after treatment is missing). Patients were instructed to keep their eyes open and visually fixate on a point marked by a grey cross (18cm wide, 18cm tall, 2cm line thickness) 110 inches in front of them (detailed information about the equipment setup and conduct with closed-loop EEG-rTMS system are available in [2, 4]). 10 of 12 SYNC and 11 of 12 UNSYNC patients completed all 30 rTMS treatments. Three sessions from three SYNC patients (one session each) were excluded from the analysis due to substantial TMS artifacts resulting from experimental equipment malfunction (Session#2 of sub-01, Session#27 of sub-05, and Session#6 of sub-08). Two patients from each treatment group missed their post-treatment scan (scan#2), so four patients' post-treatment preferred phase $\phi_{\text{post}}$ were not available. These details are listed in Table S.2.

### S.4. OFFLINE EVALUATION OF RTMS TRIGGERING ACCURACY

The method for evaluating the accuracy of the phase targeting system applied in this paper is an improvement over the method we used in our prior work, where we reported 74.4 to 95.5% of pulses were triggered within $\pm 90^\circ$ of the targeted phase [5]. Since the offline phase recovery of an EEG signal having TMS artifacts is not particularly accurate, we use the 5-min pre-treatment resting state data to investigate the phase-locked triggering accuracy in our study. Specifically, we reran the EEG-rTMS treatment system with the 5-min signals as inputs and recorded each synthetic rTMS triggering our system would choose.

These simulations were run for each SYNC subject, with one session at the beginning (S#4) and one session at the end of the treatment (S#25). These sessions were chosen to account for session variation and possible improvement of triggering accuracy (e.g., alpha oscillation becomes more stable towards the end of the sessions, resulting in better target phase estimation).

The accuracy of phase-locked rTMS triggering was evaluated after all treatment sessions were done for each patient. The EEG-rTMS system simulations were run using the same parameters (IAF and RMSE) as rTMS treatment S#4 and S#25. The system was run with the Magstim TMS device turned off, so rTMS pulse train triggers were produced and recorded with event markers in the data file without the rTMS-induced artifacts. We computed the EEG phase at the trigger event marker, compared it with the individual target phase, and obtained an estimate of triggering accuracy. The results showed no significant difference in triggering accuracy between S#4 and S#25 for the SYNC subjects, so the triggering events from the two sessions were merged to generate the triggering accuracy report (see Table S.3). The average of the error ($\phi$) across all reported SYNC subjects is $\pm 77^\circ$, which is a slight improvement compared to our previous estimates [5], though not as good as the phase-locking system introduced in Zrenner's work [6] where they only targeted two phases. Their phase estimation accuracy is $\pm 53^\circ$ if targeted at positive peak (i.e., $0^\circ$) and $\pm 55^\circ$ if targeted at negative peak (i.e., $180^\circ$).

A Wilcoxon rank sum test result between the R and NR groups revealed that the NR group

**Table S.2.** Summary of EEG-rTMS experiment and stimulation parameters. Column *Sub ID* is the subject ID for each participant. Column *Group* shows which experiment group they belong to (SYNC/UNSYNC). Column *R/NR* indicates the clinical outcome (R/NR). Column *# of Sessions* shows how many sessions have been completed and included in the analysis. Column *IAF* is the individual alpha frequency across treatments ($\mu \pm \sigma$). Column *RMSE* is the root mean square error across treatments ($\mu \pm \sigma$). Column $\phi_{pre}$, $\phi_{post}$ and $\phi_{ent}$ represents individual's pre-treatment preferred phase, post-treatment preferred phase, and entrainment phase, respectively.

| Sub ID | Group | R/NR | # of Sessions | IAF (Hz) | RMSE | $\phi_{\text{pre}}$ | $\phi_{\text{post}}$ | $\phi_{\text{ent}}$ |
|---|---|---|---|---|---|---|---|---|
| sub-01 | SYNC | R | 28/29 | $8.2 \pm 1.9$ | $0.09 \pm 0.03$ | 64° | 292° | 223° |
| sub-02 | SYNC | NR | 30/30 | $8.5 \pm 0.7$ | $0.28 \pm 0.11$ | 29° | 84° | 315° |
| sub-03 | SYNC | NR | 27/27 | $7.9 \pm 0.8$ | $0.15 \pm 0.02$ | 177° | 4° | 337° |
| sub-04 | SYNC | R | 30/30 | $10.8 \pm 0.8$ | $0.20 \pm 0.07$ | 23° | 22° | 271° |
| sub-05 | SYNC | R | 29/30 | $6.9 \pm 0.5$ | $0.16 \pm 0.02$ | 174° | 207° | 288° |
| sub-06 | SYNC | NR | 30/30 | $7.0 \pm 0.5$ | $0.25 \pm 0.05$ | 252° | 181° | 139° |
| sub-07 | SYNC | R | 30/30 | $10.7 \pm 0.8$ | $0.17 \pm 0.03$ | 355° | 178° | 189° |
| sub-08 | SYNC | NR | 29/30 | $10.9 \pm 1.8$ | $0.08 \pm 0.01$ | 313° | 34° | 339° |
| sub-09 | SYNC | NR | 30/30 | $9.6 \pm 0.7$ | $0.15 \pm 0.07$ | 159° | – | 81° |
| sub-10 | SYNC | R | 30/30 | $10.4 \pm 0.3$ | $0.44 \pm 0.05$ | 192° | 103° | 256° |
| sub-11 | SYNC | R | 30/30 | $6.9 \pm 0.2$ | $0.28 \pm 0.08$ | 222° | 105° | 253° |
| sub-12 | SYNC | NR | 30/30 | $9.2 \pm 2.9$ | $0.29 \pm 0.11$ | 46° | – | 283° |
| sub-13 | UNSYNC | NR | 30/30 | $8.3 \pm 0.6$ | $0.16 \pm 0.02$ | 313° | 311° | 210° |
| sub-14 | UNSYNC | NR | 30/30 | $11.1 \pm 0.9$ | $0.38 \pm 0.05$ | 302° | 184° | 285° |
| sub-15 | UNSYNC | R | 30/30 | $9.7 \pm 1.3$ | $0.16 \pm 0.05$ | 34° | 242° | 261° |
| sub-16 | UNSYNC | R | 30/30 | $7.0 \pm 0.4$ | $0.17 \pm 0.08$ | 299° | – | 204° |
| sub-17 | UNSYNC | R | 30/30 | $8.3 \pm 0.5$ | $0.16 \pm 0.04$ | 288° | 165° | 12° |
| sub-18 | UNSYNC | R | 28/28 | $7.3 \pm 1.2$ | $0.19 \pm 0.03$ | 315° | 90° | 0° |
| sub-19 | UNSYNC | NR | 30/30 | $11.1 \pm 0.8$ | $0.15 \pm 0.08$ | 207° | 28° | 203° |
| sub-20 | UNSYNC | R | 30/30 | $8.4 \pm 0.6$ | $0.15 \pm 0.03$ | 293° | 125° | 293° |
| sub-21 | UNSYNC | R | 30/30 | $9.5 \pm 0.4$ | $0.17 \pm 0.03$ | 271° | – | 290° |
| sub-22 | UNSYNC | R | 30/30 | $7.6 \pm 0.3$ | $0.18 \pm 0.05$ | 238° | 266° | 265° |
| sub-23 | UNSYNC | NR | 30/30 | $10.3 \pm 0.6$ | $0.21 \pm 0.04$ | 18° | 36° | 225° |
| sub-24 | UNSYNC | NR | 30/30 | $10.3 \pm 1.7$ | $0.11 \pm 0.01$ | 45° | 71° | 245° |

demonstrated higher triggering accuracy than the R group (two-tail: $p = 0.011$). This finding suggests the potential value of measuring the individual preferred phase during the treatment, as the group with diminished precision in triggering exhibited better clinical improvement by still aligning the rTMS pulses around their preferred phases despite the phase shift. However, it is important to note that the offline triggering accuracy, obtained through simulation using resting data, may not fully reflect the actual triggering accuracy during the experimental sessions (e.g., no significant difference in RMSE between R and NR groups). Further investigations are warranted to explore this observation.

**Table S.3.** Phase triggering accuracy for all SYNC patients. Column $\phi_{tar} = \phi_{pre}$ is the phase we targeted when the rTMS-EEG system triggered, which is equal to the pre-treatment individual preferred phase. Column *# of simulated triggers* shows the total number of trigger attempts made during the simulation on the 5-min pre-treatment resting data from two selected sessions. Column *error($\phi$)* is the average difference between the recovered and target phases.

| Sub ID | R/NR | $\phi_{\text{tar}} = \phi_{\text{pre}}$ | # of simulated triggers | error($\phi$) |
|---|---|---|---|---|
| sub-01 | R | 64° | 31 | ±74° |
| sub-02 | NR | 29° | 24 | ±74° |
| sub-03 | NR | 177° | 28 | ±77° |
| sub-04 | R | 23° | 34 | ±84° |
| sub-05 | R | 174° | 27 | ±77° |
| sub-06 | NR | 100° | 29 | ±65° |
| sub-07 | R | 355° | 34 | ±95° |
| sub-08 | NR | 313° | 31 | ±69° |
| sub-09 | NR | 159° | 32 | ±66° |
| sub-10 | R | 192° | 30 | ±89° |
| sub-11 | R | 222° | 23 | ±97° |
| sub-12 | NR | 46° | 34 | ±59° |

### S.5. GLOBAL/LOCAL MEAN FIELD POWER

#### A. Global mean field power (GMFP) at resting-state

The result of a paired t-test between GMFP measured at the 3-min resting-state before and after the treatment of each treatment session is shown in the first sub-table of Table S.4. There is a significant decrease in GMFP at weeks #1, #2, #4, and #6 within the SYNC group (the significance level was shown in Fig. 2a of the main text). No such observation is found within the UNSYNC group. Additionally, there is no significant difference between SYNC and UNSYNC groups at week#1 (see the second sub-table of Table S.4), which indicates both groups are at a similar level of global cortical excitability at the beginning of treatment. As the treatment progressed, the difference in GMFP at the 3-minute RS before the treatment between groups appeared (at week#2, #4, and #6) and decreased (week#2) or disappeared (week#4 and #6) after the treatment. This observation suggests that this synchronization-induced session-level modulation of GMFP might not be sustainable for an extended period (i.e., the subject who obtained low GMFP after treatment this week might come in with a high GMFP for the following week). For the overall decrease of GMFP observed over 6-week treatment within the UNSYNC group, it might be more sustainable (although no significant paired decrease), which might help us explain the clinical improvement within the UNSYNC group (see section B for more details).

**Table S.4.** Paired t-test results between GMFP measured at the 3-min resting state before and after each treatment session within each group and unpaired t-test results between SYNC and UNSYNC groups within each RS period. The significance level of the test result was indicated by a black asterisk, where (***) indicates significance under a 99.9% confidence level, (**) indicates significance under a 99% confidence level, and (*) indicates significance under a 95% confidence level.

| | SYNC | UNSYNC | RS (before) | RS (after) |
|---|---|---|---|---|
| Week | pre vs post | pre vs post | SYNC vs UNSYNC | SYNC vs UNSYNC |
| #1 | 0.029(*) | 0.770 | 0.630 | 0.146 |
| #2 | 0.034(*) | 0.634 | 0.001(**) | 0.022(*) |
| #3 | 0.067 | 0.547 | 0.154 | 0.317 |
| #4 | <0.001(***) | 0.466 | 0.009(**) | 0.452 |
| #5 | 0.318 | 0.652 | 0.147 | 0.484 |
| #6 | 0.040(*) | 0.642 | 0.024(*) | 0.363 |

### B. Generalized Linear Mixed-effects Model (GLMM) with GMFP

The main text discussed the correlation between GMFP and Hamilton Depression Rating Scale (HDRS) changes with weekly and group comparisons. Here we investigated the relationship between GMFP and HDRS changes with a GLMM model [7]. We used the GLMM in Matlab (Statistics and Machine Learning Toolbox, Matlab 2018b, Mathworks, USA) to analyze changes in HDRS across weeks as a function of GMFP. A GLMM is an extension to the generalized linear model (GLM) in which the linear predictor contains random effects in addition to the fixed effects [8]. The fixed effects in the model included GMFP measured at the resting state after treatment, changes in GMFP at the resting state within a session, treatment week number, and the interaction between the GMFP changes and treatment week number. The subject difference was modeled by grouping variable sub as random effects. Therefore, the final model is:

$$HDRS \sim 1 + GMFP_{\text{after}} + \Delta GMFP * week + (1|sub) + \epsilon \tag{S1}$$

where $HDRS$ is the HDRS measured weekly prior to the commencement of treatment for the following week; $GMFP_{\text{after}}$ is the weekly average of GMFP measured at the resting state after each treatment; $\Delta GMFP$ is the weekly average of changes in GMFP at the resting state within a session; $week$ is the number of treatment weeks (i.e., $1, 2, \cdots, 6$); $sub$ represents each subject/patient (i.e., $1, 2, \cdots, 23$, one UNSYNC subject is excluded because of missing after treatment resting-state recording); $\epsilon$ is the model residual.

The fixed effects of the GLMM are listed in Table S.5 and the results of the random effects are not reported here because no significant subject-level difference was found. As reported in the main text, there is a significant positive correlation between $HDRS$ and $\Delta GMFP$ regardless of treatment group-i.e. the greater a decrease in GMFP after each treatment session, the smaller the HDRS and better clinical improvement ($p = 0.019$). Additionally, the effect of $\Delta GMFP$ decreases with the increase of treatment week ($p = 0.002$). $GMFP_{\text{after}}$ is a significant factor that positively associates with changes in $HDRS$ as well. The greater/smaller GMFP measured at the resting state after each treatment, the greater/smaller HDRS would be. This finding further confirmed the observation mentioned in section A that although there is a session-level modulation of GMFP with the EEG-synchronized rTMS treatment, maintaining a low global cortical excitability after treatment might be more important than showing a continuous decrease in excitability. It is the reason why when we constructed the excitability index (EI1) in Fig. 6a1, we prioritized the comparison between $GMFP_{\text{after}}$ and $GMFP_{\text{thres}}$ before the comparison between $\Delta GMFP$ and 0.

**Table S.5.** Fixed effects coefficients of GLMM (95% CIs).

| Name | Estimate | DF | $p$-value | Lower CI | Upper CI |
|---|---|---|---|---|---|
| (Intercept) | 25.614 | 133 | <0.001(***) | 22.173 | 29.055 |
| $\Delta GMFP$ | 6.456 | 133 | 0.019(*) | 1.074 | 11.838 |
| $week$ | -2.020 | 133 | <0.001(***) | -2.303 | -1.738 |
| $GMFP_{\text{after}}$ | 4.021 | 133 | 0.047(*) | 0.055 | 7.987 |
| $\Delta GMFP : week$ | -2.161 | 133 | 0.002(**) | -3.522 | -0.801 |

#### C. Local mean field power (LMFP) at resting state

LMFP was calculated at four ROIs based on our previous findings [2]. The scalp map in Figure S.4 defines the ROIs and the electrodes used in the analysis – i.e., near target region (tar: FP1, F3, and F7), contralateral to the target region (con: FP2, F4, and F8), media-frontal region (med: FZ, FC1, and FC2) and occipital region (occ: O1, Oz, and O2). The comparison of LMFP between groups was performed similarly to what was done for GMFP. Given results in Table S.6, we found that session-level synchronized rTMS-induced excitability decrease was strongest in the occipital region where there is a significant decrease of LMFP at the resting state (occ>con>tar>med, based on how many weeks has significant LMFP decrease). Moreover, the session-level changes observed in the occipital region were more sustainable compared to other ROIs since the LMFP of both groups at the resting state before treatment was at a similar level (except week#4 and #6). In general, our findings are consistent with other research with MDD populations, which found reduced activation in the frontal lobe, parietal lobe, and occipital regions in patients vs health controls, when subjects were presented positive and neutral pictures [9–12]. However, additional source-level analysis is needed to confirm the source regions of those changes; with the limited number of EEG channels we have, this is not achievable in our study.

#### D. Global/Local mean field power at the inter-pulse train interval

We also computed the GMFP and LMFP changes at the IPI period to investigate the immediate excitability changes after the rTMS pulse train. As introduced in the Method section of the main text, ten temporal windows were selected: -2500 to -2000 ms (-T5), -2000 to -1500 ms (-T4), -1500 to

**Table S.6.** Paired t-test results between LMFP measured at the 3-min resting state before and after each treatment session within each group and unpaired t-test results between SYNC and UNSYNC groups within each RS period. Four ROIs were selected, which are near the target region (tar), contralateral to the target region (con), media-frontal region (med), and occipital region (occ), respectively. Significant paired t-test indicates pre>post, and significant unpaired t-test indicates SYNC>UNSYNC.

| ROI | Week | SYNC before vs after | UNSYNC before vs after | RS (before) SYNC vs UNSYNC | RS (after) SYNC vs UNSYNC |
|---|---|---|---|---|---|
| tar | #1 | 0.268 | 0.694 | 0.136 | 0.331 |
| | #2 | 0.285 | 0.332 | 0.005(**) | 0.139 |
| | #3 | 0.089 | 0.884 | 0.016(*) | 0.142 |
| | #4 | 0.001(**) | 0.123 | 0.024(*) | 0.023(*) |
| | #5 | 0.730 | 0.207 | 0.148 | 0.137 |
| | #6 | 0.013(*) | 0.930 | 0.004(**) | 0.002(**) |
| con | #1 | 0.058 | 0.142 | 0.304 | 0.712 |
| | #2 | 0.011(*) | 0.145 | 0.002(**) | <0.001(***) |
| | #3 | 0.050 | 0.192 | 0.147 | 0.093 |
| | #4 | <0.001(***) | 0.992 | 0.006(**) | 0.786 |
| | #5 | 0.878 | 0.667 | 0.045(*) | 0.311 |
| | #6 | 0.018(*) | 0.745 | 0.002 (**) | 0.006(*) |
| med | #1 | 0.888 | 0.106 | 0.319 | 0.823 |
| | #2 | 0.084 | 0.646 | 0.218 | 0.979 |
| | #3 | 0.559 | 0.331 | 0.190 | 0.142 |
| | #4 | 0.008(**) | 0.334 | 0.028(*) | 0.399 |
| | #5 | 0.890 | 0.506 | 0.707 | 0.947 |
| | #6 | 0.412 | 0.552 | 0.004(**) | 0.024(*) |
| occ | #1 | 0.013(*) | 0.155 | 0.793 | 0.241 |
| | #2 | 0.007(**) | 0.398 | 0.139 | 0.738 |
| | #3 | 0.005(**) | 0.303 | 0.295 | 0.365 |
| | #4 | 0.004(**) | 0.041(*) | 0.041(*) | 0.212 |
| | #5 | 0.028(*) | 0.270 | 0.418 | 0.998 |
| | #6 | 0.006(**) | 0.347 | 0.007(**) | 0.425 |

-1000 ms (-T3), –1000 to -500 ms (-T2), –500 to -128 ms (-T1), 128 to 500 ms (T1), 500 to 1000 ms (T2), 1000 to 1500 ms (T3), 1500 to 2000 ms (T4) and 2000 to 2500 ms (T5), with 0 ms corresponding to the entire rTMS pulse train. The first 128ms after and last 128ms before the rTMS pulse train were not analyzed due to filtering artifacts caused by zero padding at the beginning and end of the epoch (marked as the shadow area in Figure S.1a.2). The filter order and the sampling rate determined the length of this time window. For SYNC subjects, global cortical excitability in the IAF (6 to 13 Hz) band represented by GMFP shows a significantly immediate increase after the rTMS pulse train (i.e., T1: 128 ms to 500 ms, see Figure S.1 a.1) compare to the 2.5 seconds pre-stimulation baseline. The SYNC group also had higher post-stimulation excitability than the UNSYNC group at T1. Counter-intuitively, the global cortical excitability of both groups decreases to a similar level below their pre-stimulation baseline and is maintained low for at least 1000 msecs (during the T2 and T3 period). The continuous excitability changes within the time period -T1 and T1 for both groups are shown in Figure S.1 a.2. We can observe that there is an approximately 25 ms constant delay (P1:$\Delta t = 20$ ms, P2: $\Delta t = 36$ ms and P3: $\Delta t = 20$ ms) of the appearance of the peak in the UNSYNC group compared to the SYNC group. The average event-related potentials (ERPs) of each group at each peak moment (first three peaks after stimulation) are presented in Figure S.1 a.3. Based on the LMFP calculated in the four selected ROIs, the immediate increase in global cortical excitability is likely contributed by the near target and contralateral to target regions. None of these selected ROIs is an obvious primary contributing source for the subsequent decrease in global cortical excitability, based on their LMFP changes (see Figure S.1b).

### S.6. WEIGHTED INTER-TRIAL PHASE COHERENCE (WITPC)

#### A. Calculation of wITPC and $\phi_{\text{ent}}$

Since the accuracy of the phase estimation by the Hilbert transform on a specific frequency band is dependent on that band's signal-to-noise ratio (SNR) [13, 14], we updated the ITPC calculation that takes the arithmetic mean of all trials to the power-weighted mean (i.e., wITPC) to consider the ratio of the quasi-alpha (6–13 Hz) wave to other EEG components (1–30 Hz). Therefore, relative power was defined as the ratio of absolute quasi-alpha power to the total power calculated from 1 to 30 Hz (spanning delta, theta, alpha and beta bands). 6 to 13 Hz was used because it was the range used to identify the IAF for each subject during the rTMS triggering. The trial weight was determined by its relative quasi-alpha power before the TMS pulse train delivering (see equation S2 and S3), and the Hilbert Transform was applied to the post-stimulation period to estimate the instantaneous phase $\phi_{N,j}(t)$ of EEG signal $x_{N,j}(t)$ locked to the last TMS pulse for trial $N$ and channel $j$ (see Figure S.2). Lastly, the wITPC for the epoched post rTMS pulse train period (i.e., $t \in [0, 2.5]$s) is calculated with equation S4 (same as equation (4) in the main text. The details of wITPC calculation can also be found in [2, 4]).

$$\overline{\alpha}_N = \frac{\int_6^{13} P_{N,f}^{\text{tar}} df}{\int_1^{30} P_{N,f}^{\text{tar}} df} = \frac{\int_6^{13} \frac{1}{3}(P_{N,f}^{\text{FP1}} + P_{N,f}^{\text{F3}} + P_{N,f}^{\text{F7}}) df}{\int_1^{30} P_{N,f}^{\text{tar}} df} \tag{S2}$$

$$\omega_N = \frac{\overline{\alpha}_N}{\sum_{N=1}^{N=75} \overline{\alpha}_N} \tag{S3}$$

where $\int_{f_1}^{f_2} P_{N,f}^{j} df$ is the integral of power between $f_1$ and $f_2$ of channel $j$ for trial $N$; tar refers to the near target region which includes electrode FP1, F3, F7, so $j = \{\text{FP1, F3, F7}\}$; $\overline{\alpha}_N$ is the relative

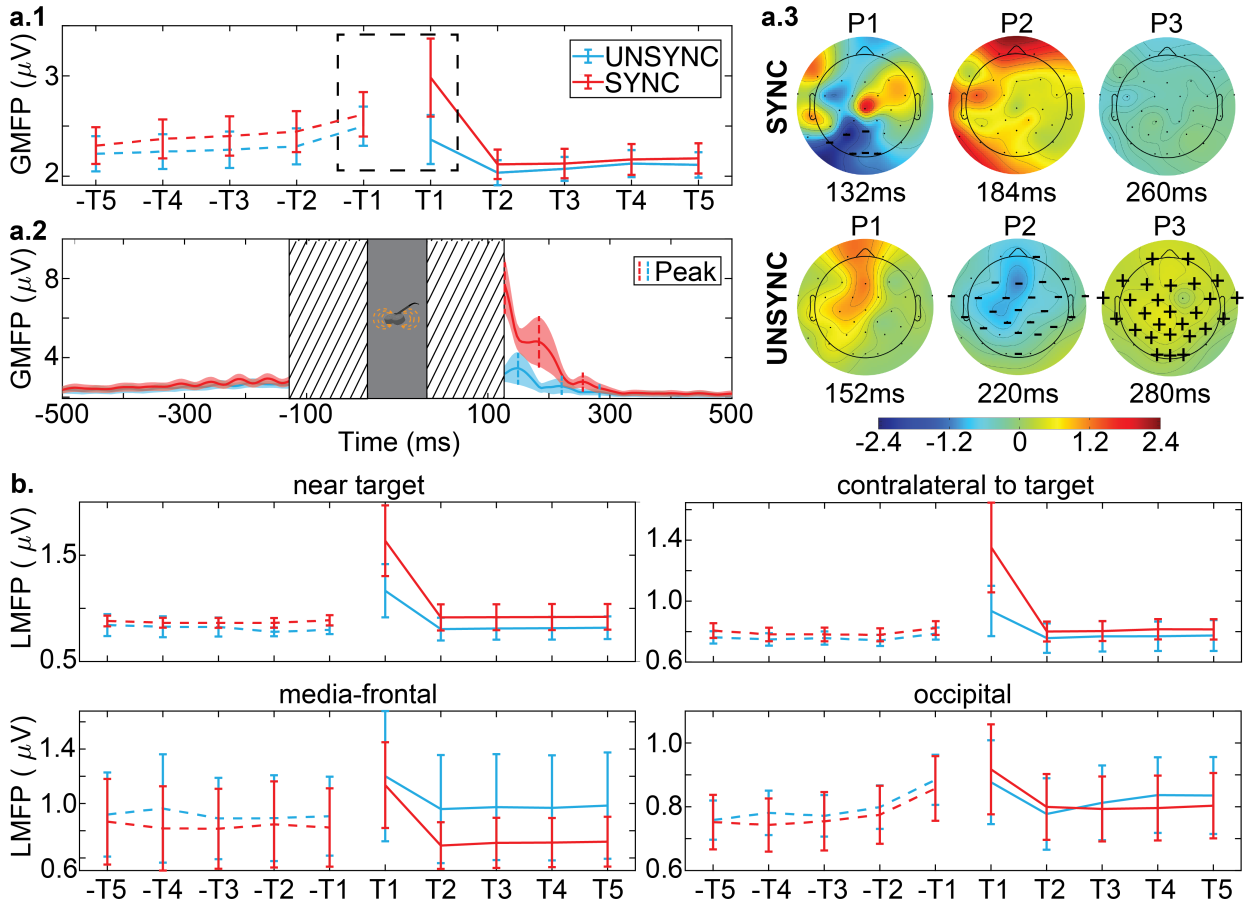


**Fig. S.1.** GMFP/LMFP measured at the inter-pulse train interval. **a.1**. GMFP measured at the selected ten temporal windows. There is a significant difference in global cortical excitability between SYNC and UNSYNC groups at T1 ($p < 0.001$(***)). **a.2** A zoomed-in version of GMFP changes within the temporal window -T1 and T1 where continuous changes were shown. The filter effect was marked as the shadow area, and the rTMS pulse train delivering period was highlighted as a gray rectangular area. After an immediate increase, the global cortical excitability maintains a lower level since ∼280 ms. The first three excitability peaks after stimulation were detected for both groups to illustrate their corresponding ERPs, and there is an approximately 25 ms constant delay of peak appearance in the UNSYNC group. **a.3**. Each group's average event-related potentials (ERPs) at each peak moment. Within the SYNC group, there is a significant decrease in the ERPs observed at the occipital region at 132 ms after the rTMS stimulation. For the UNSYNC group, there is a significant decrease of ERPs observed at most electrodes (except FP1 and F7 at the left pre-frontal and some electrodes at the parietal such as P3 and P7) at 220 ms after the rTMS stimulation and a significant increase of ERPs at 280 ms (multi-comparison corrected, an electrode is marked with a "**-**" sign if there is a significant decrease with $p < 0.05$ and a "**+**" sign if there is a significant increase with $p < 0.05$). **b**. LMFP was measured at the selected ten temporal windows. The LMFP measured at the near target and contralateral to target region contribute the most to the immediate GMFP increase after rTMS within the SYNC subjects (i.e., the difference observed at T1, SYNC>UNSYNC (tar): $p = 0.004$, SYNC>UNSYNC (con): $p = 0.002$).

alpha power for trial $N$ of each session; $\omega_N$ is the trial weight for trial $N$ of each session.

$$wITPC(t) = |\frac{1}{3}\sum_{j=1}^{j=3}\sum_{N=1}^{N=75} e^{i\times\phi_{N,j}(t)} \times \omega_N| \tag{S4}$$

#### B. Definition of the first post-stimulation wITPC peak

The brain synchronization represented by phase entrainment in the quasi-alpha band in our study is defined as the level of phase alignment in the post-TMS stimulation EEG across all trials within each session, as measured by wITPC. The larger the wITPC value, the more consistent the EEG phase across trials after the rTMS pulse train has ended. We define the first post-stimulation wITPC peak (wITPC$^{[1]}$) as the first local maximum of the wITPC following the last TMS pulse in a pulse train (see Figure S.2). The details of wITPC$^{[1]}$ detection is available in S.6 of [4]. To further confirm that the phase entrainment we detected after rTMS is not a result of phase-locking itself, Fig S.3a shows the summary statistics of the time (unit: ms) when the first peak is detected. All subjects have varied detected peak times (mostly between 200 to 400 ms) across entire treatment sessions, which indicates that the entrainment is not a result of phase-locking itself but a TMS-induced phenomenon.

#### C. GLMM with wITPC$^{[1]}$

In our preliminary analysis with 15 subjects, we found that wITPC$^{[1]}$ increased across sessions only when rTMS was synchronized to individual preferred phase [2]. The updated GLMM results with 24 subjects are consistent with our previous finding [15]. Specifically, we observed a statistically significant effect of the interaction between the factors session-number (1–30) and treatment group (SYNC and UNSYNC) on wITPC$^{[1]}$ as the fixed effect of a GLMM at the near target region (GLMM used here is the same one as introduced in [2], see equation S5; we reported the results at Table S.7: $\beta = 0.013$, $p = 0.002$).

$$\ln\frac{\text{wITPC}^{[1]}}{1-\text{wITPC}^{[1]}} \sim stimf + (session + \bar{\alpha}_P) * condition + (1|sub) + \epsilon, \ln\frac{\text{wITPC}^{[1]}}{1-\text{wITPC}^{[1]}} \in [0,1] \tag{S5}$$

where wITPC$^{[1]}$ refers to the first post-stimulation wITPC peak value for each session; $stimf$ refers to the stimulation frequency for each session (i.e., IAF); $session$ is the corresponding session number (i.e., 1, 2, $\cdots$, 30); $\bar{\alpha}_P$ is the relative quasi-alpha power for each session; $condition$ is either SYNC(1) or UNSYNC(-1); $sub$ represents each subject (i.e., 1, 2, $\cdots$, 24). In addition, because the range of wITPC$^{[1]}$ is between 0 and 1, the logit link function is applied in this linear model.

Interestingly, compared to our previous result, there is a change in the significance level of stimulation frequency ($\beta = 0.037$, $p = 0.020$) and relative quasi-alpha power ($\beta = -1.318$, $p = 0.005$), which indicates a positive correlation between the stimulation frequency (i.e., IAF) and the wITPC$^{[1]}$ and negative correlation between relative quasi-alpha power with wITPC$^{[1]}$ (note: no significant difference between groups in stimulation frequency and relative quasi-alpha power). These results suggest how the changes of wITPC$^{[1]}$ – phase synchronization – would be related to the dominant frequency and its power. For example, it might be harder to synchronize the signal when relative alpha power is high (where the alpha band is dominant among other frequency bands). Further investigation of this observation would help us understand how to increase the chances of inducing phase synchronization.

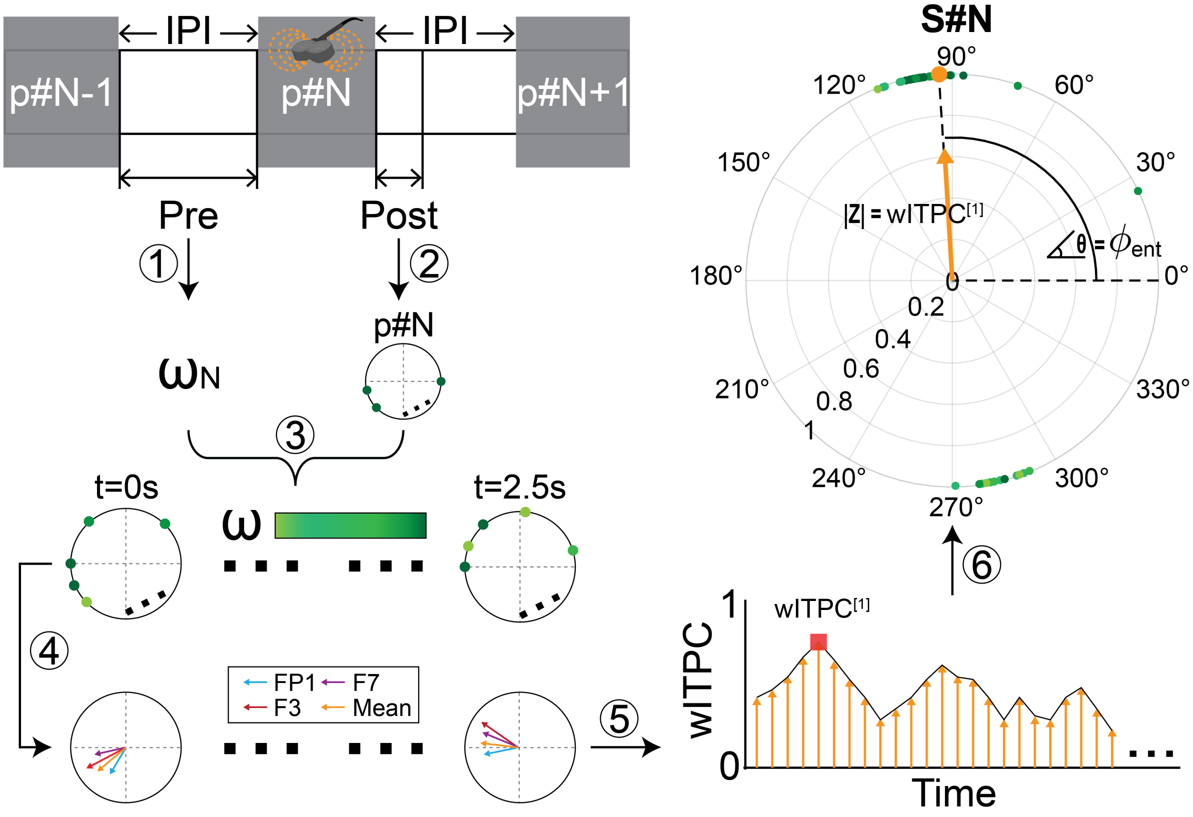


**Fig. S.2.** Flowchart of calculating wITPC[1] and $\phi_{\text{ent}}$ within each session. Each black arrow indicates the processing flow, starting at the upper left and going counterclockwise to the upper right. First, two datasets were epoched. One is referred to as "Pre" and the other as "Post" based on each TMS pulse train. The "Pre" data was used to calculate trial weights, and the "Post" data was used to obtain the phase. ① For pulse train p#$N$, the trial weight, $\omega_N$, was calculated based on the relative alpha power measured during the "Pre" segment. ② The phase of the "Post" segment was obtained by the Hilbert transform and shown in polar coordinates by applying Euler's formula. ③ Repeat the process outlined in the previous two steps for all the pulse trains within one session. It can generate polar coordinates plots sequentially encompass all the trials over time (i.e., $t = 0$s to $t = 2.5$s after the rTMS pulse train). The illustration figure shows all trials at the same post-stimulation time . Each dot indicates one trial. Darker green means greater $\omega$, while lighter green means smaller $\omega$. ④ The corresponding trial average (with power weights $\omega$) at each time point was calculated with eq (6), resulting in a vector where the magnitude indicates the phase coherence across time for different trials and the angle indicates the direction of the phase alignment. The wITPC was calculated for each electrode with one ROI. The near target region was shown as an example which includes electrodes FP1 (blue), F3 (red), and F7 (purple), and the mean (orange) of the ITPC was also calculated across the three electrodes. ⑤ The magnitudes of the ROI vector were plotted by time, and the moment where the first magnitude peak (wITPC[1]) occurs was taken to present the post-stimulation phase synchronization for that session. ⑥ Based on the selected time moment (i.e., when the wITPC[1] detected), the corresponding angle value of that vector is defined as $\phi_{\text{ent}}$. The illustration figure shows the wITPC[1] and $\phi_{\text{ent}}$ of one section as an example, and same procedures are repeated for all treatment sessions.

**Table S.7.** Fixed effects coefficients of GLMM (95% CIs).

| Name | Estimate | DF | $p$-value | Lower CI | Upper CI |
|---|---|---|---|---|---|
| (Intercept) | -0.648 | 704 | 0.009(**) | -1.135 | -0.162 |
| $stimf$ | 0.037 | 704 | 0.020(*) | 0.0058 | 0.069 |
| $\bar{\alpha}_P$ | -1.318 | 704 | 0.005(**) | -2.230 | -0.405 |
| $session$ | 0.004 | 704 | 0.157 | -0.002 | 0.010 |
| $condition$ | -0.010 | 704 | 0.970 | -0.517 | 0.498 |
| $\bar{\alpha}_P : condition$ | -0.363 | 704 | 0.549 | -1.551 | 0.826 |
| $session : condition$ | 0.013 | 704 | 0.002(**) | 0.005 | 0.022 |

### S.7. ENTRAINMENT PHASE

Different from our hypothesis, another common hypothesis is that there is a generally preferred stimulation phase common across all individuals – e.g., negative peak ('trough' of oscillation): $\frac{3\pi}{2}$) that would optimize the neurostimulation efficacy [6, 16]. In Table S.2, the summary of pre-treatment preferred phase ($\phi_{\text{pre}}$), pre-treatment preferred phase ($\phi_{\text{post}}$) and entrainment phase ($\phi_{\text{ent}}$) of each subject is shown. There is no significant difference in $\phi_{\text{pre}}$ between R and NR within the SYNC and UNSYNC group, but we found that the majority of the R group within the UNSYNC group has their $\phi_{\text{pre}}$ located around the negative peak (Rao's spacing test[1]: $p = 0.050$) which might partially explain why the unsynchronized rTMS simulation was efficacious for this group. Additionally, the fact that the majority of the R (repsonders) within the SYNC group had their $\phi_{\text{ent}}$ located around the negative peak ($p = 0.050$) provides further evidence for the significance of the negative peak in neurostimulation (see Fig. S.3b). Conversely, the majority of the NR group within the UNSYNC group had their $\phi_{\text{ent}}$ located around the negative peak (Rao's spacing test: $p = 0.050$), which suggests that the direction of phase entrainment might only matter when the synchronized stimulation is applied. This observation might be why the treatment efficacy/clinical improvement is more predictable by biomarker changes within the SYNC group.

### S.8. PERMUTATION TEST OF THE ENTRAINMENT OBSERVED WITHIN SYNC GROUP

Within all twelve SYNC subjects, we randomly re-assigned the responder and non-responder labels to all SYNC subjects. We checked whether the new distribution of the responder/non-responder (R/NR) label within all SYNC subjects would still be similar to have the same separation between responder and non-responder–i.e., all clinical responders within the SYNC group have higher entrainment as shown in the Fig. 3 of main text. Repeating this process 1000 times, around 3% of the result would be the same distribution as our founding even with allowing one SYNC responder labeled as non-responder (making the separation between R and NR less clear).

### S.9. SEPARATION BETWEEN EXCITABILITY AND ENTRAINMENT

In the Fig. 4 of main text, we showed the separation observed in EI (relevant to ΔGMFP1) and EI2 (relevant to $\phi_{\text{ent}}$) by groups (SYNC:R, SYNC:NR, UNSYNC:R, and UNSYNC:NR). As the figure

[1]Rao's spacing test by comparing distances between points on a circle to those expected from a uniform distribution [17].

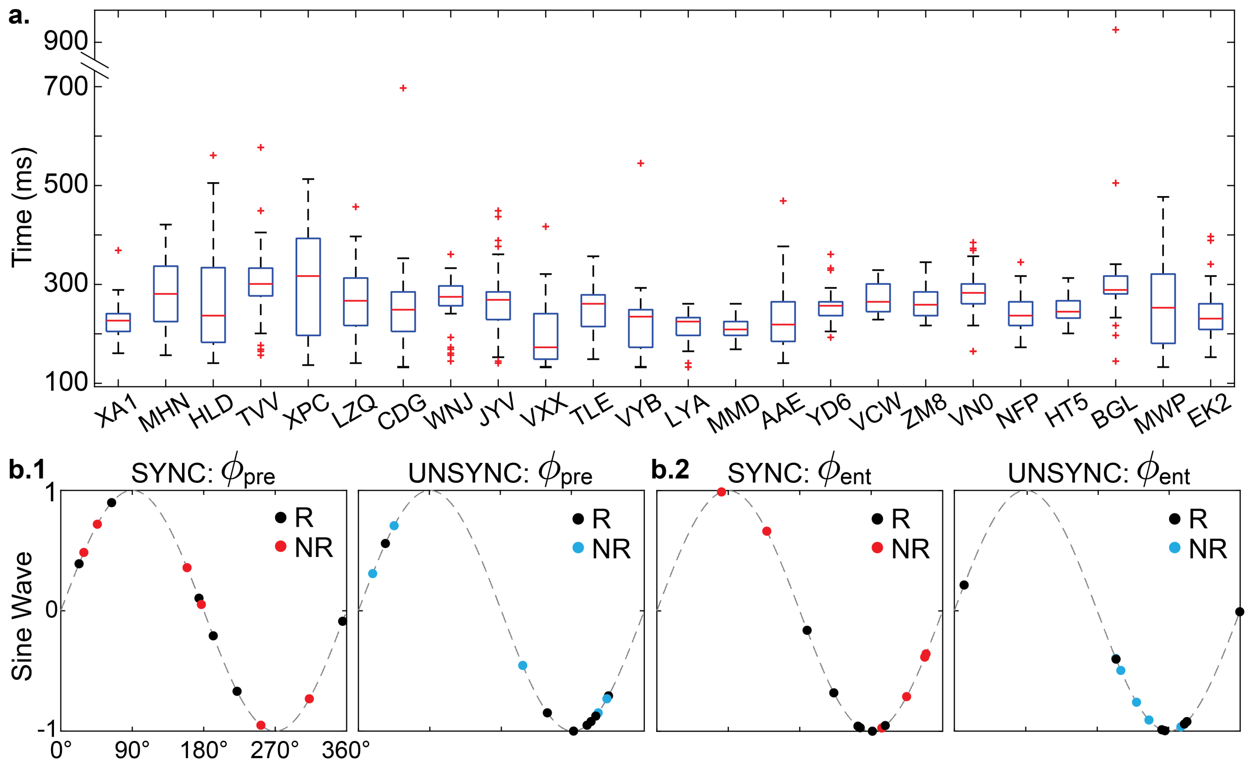


**Fig. S.3.** Distribution of the peak detection time of wITPC[1], pre-treatment phase $\phi_{pre}$ and entrainment phase $\phi_{ent}$. **a.** Distribution of the peak detection time of wITPC[1]. No significant difference in the peak time between treatment groups or between R and NR. All subjects have varied first peak times (mostly 200 to 400 ms) across treatment sessions. **b.1** Distribution of $\phi_{pre}$ in sine wave. No significant difference between treatment groups or between R and NR, but the $\phi_{pre}$ of R within the UNSYNC group are not uniformly distributed (Rao's spacing test: $p = 0.050$, located around the 'trough'). **b.2** Distribution of $\phi_{ent}$ in sine wave. No significant difference between treatment groups or between R and NR. However, the $\phi_{ent}$ of R within the SYNC group and NR within the UNSYNC group are not uniformly distributed (Rao's spacing test: $p = 0.050$, located around the 'trough').

presents, the clinical responders (R) within the SYNC group have the greatest excitability decrease and the highest entrainment compared to the other subgroups. The results of statistical comparison between groups in excitability (represented by ΔGMFP) are reported in Fig. 2a of the main text. The results of entrainment (represented by $\Delta SD(\phi) \times$ # of entrained sessions) are available in Table S.8. Because entrainment is measured based on all treatment sessions, each subject only has one measurement. We used the Mann-Whitney-Wilcoxon test instead for comparison with small sample sizes [18].

### S.10. PHASE-LOCKING VALUE (PLV)

Surrogate data testing was applied on the phase-locking value calculated at the resting state [19, 20]. Since no session-level increase or decrease in PLV was found within either group, the average PLV across all treatment sessions was used for comparison. For the comparison across all patients, we found that there is a significant difference between SYNC and UNSYNC groups in the PLV calculated at the resting state before and after treatment between the occipital and other three ROIs

**Table S.8.** Mann-Whitney-Wilcoxon test results of entrainment (represented by $\Delta SD(\phi) \times$ # of entrained sessions) between groups. Six subgroups are included here based on the treatment arms (SYNC and UNSYNC) and the clinic outcomes within each condition (SYNC:R, SYNC:NR, UNSYNC:R, and UNSYNC:NR).

| | Entrainment: $\Delta SD(\phi) \times$ # of entrained sessions | | | | | |
|---|---|---|---|---|---|---|
| | SYNC $N = 12$ | UNSYNC $N = 12$ | SYNC:R $N = 6$ | SYNC:NR $N = 6$ | UNSYNC:R $N = 7$ | UNSYNC:NR $N = 5$ |
| SYNC | - | 0.044(*) | - | | | |
| UNSYNC | 0.044(*) | - | - | | | |
| SYNC:R | - | - | - | 0.008(**) | 0.001(**) | 0.023(*) |
| SYNC:NR | | | 0.008(**) | - | 0.413 | 0.279 |
| UNSYNC:R | | | 0.001(**) | 0.413 | - | 0.068 |
| UNSYNC:NR | | | 0.023(*) | 0.279 | 0.068 | - |

(see Figure S.4a.1). The corresponding paired t-test results within a group were shown on the right. We can see that within the UNSYNC group, there is a significant increase of PLV among all ROI pairs after each treatment session (see Figure S.4b.1). Additionally, for the comparison between R and NR within the SYNC group, we found that clinic responders maintain at a lower PLV for all paired ROIs (except 'tar-con') compared to the clinic non-responders and no significant PLV changes after each treatment session has been observed within the R group. Surprisingly, although there is no significant difference between R and NR within the UNSYNC group, there is a small but significant PLV increase after treatment for the responders.

Moreover, clinic responders from both SYNC and UNSYNC groups have a similarly low PLV before and after treatment compared to the non-responders, which might partially help us explain why we did not observe a significant general clinic improvement difference between the two treatment groups (UNSYNC group might be easier to respond because their low synchronization across brain regions before treatment). More importantly, from the PLV comparisons, we found that the EEG-synchronized and unsynchronized rTMS moved the direction of synchronization differently, and some of the findings are consistent with other literature. For example, it is widely believed that the function of the occipital lobe is linked to the symptoms of depressed patients [11, 12], and our EEG-synchronized and unsynchronized rTMS treatment stimulates at DLPFC can affect the oscillation synchronization between occipital and other regions.

### S.11. ENTRAINMENT EFFECT AND FLIPPING EFFECT

To track the level of phase synchronization by treatment week, we developed a measure that reflects the weekly changes in entrainment phase $\phi_{ent}$. This measurement comprises two effects – the entrainment effect and the flipping effect – where we can track the phase synchronization by analyzing weekly clustering outcomes. The flowchart of the measurement is shown in Fig S.5. After each week's treatment, circular clustering with $K = 2$ was performed on all sessions' $\phi_{ent}$ until the current treatment week (e.g., after week#2, $\phi_{ent}$ of 10 sessions were included for clustering, see Fig S.5b). By comparing the difference of $\phi_{ent}$ distribution between the current treatment week

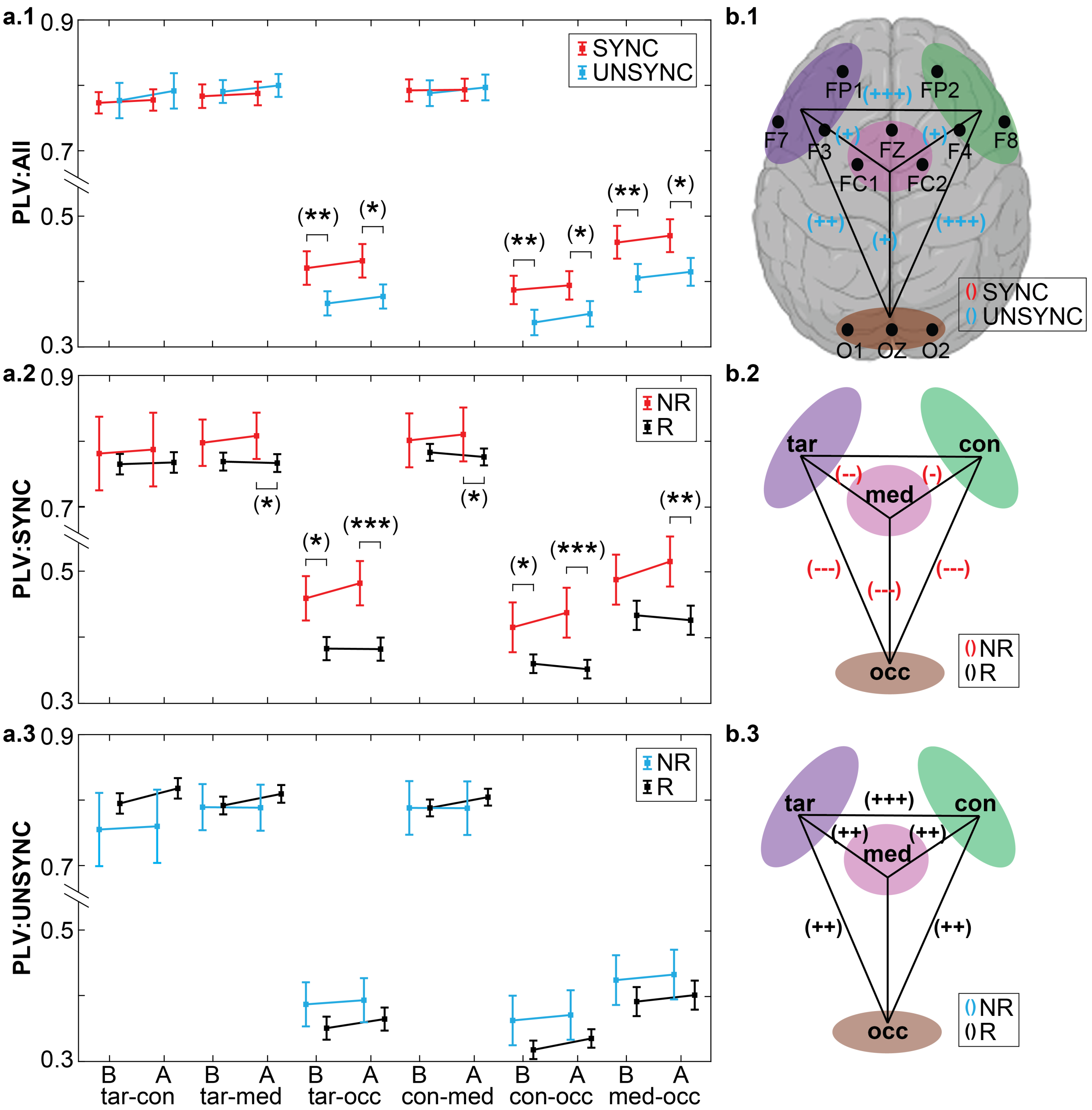


**Fig. S.4.** Comparison of PLV computed at the resting-state. **a.1** The results of unpaired t-test between SYNC and UNSYNC group were shown. Six ROI pairs are included, which are near target vs contralateral to target (tar-con), near target vs media-frontal (tar-med), near target vs occipital (tar-occ), contralateral to target vs media-frontal (con-med), contralateral to target vs occipital (con-occ), and media-frontal vs occipital (med-occ). ROIs were selected based on our research interests [2]. **b.1** The results of paired t-test on PLV changes between before(B) and after(A) each treatment within each treatment group (SYNC and UNSYNC) were presented. Results reported are multi-comparison corrected. ROI pairs marked with "**-**" sign indicates there is a significant decrease and "**+**" sign indicates there is a significant increase (significance level was indicated by the number of signs). The definitions of ROIs and its included electrodes were shown in the scalp map. Subplot **a.2** and **a.3** are similar to **a.1**, except that the comparison is performed for R and NR groups within each treatment group. Subplot **b.2** and **b.3** mirror **b.1**.

and the previous week, we can know how many sessions are in the entrained class and whether they locate in a narrower direction (i.e., entrainment effect). In addition, we measure how many sessions change their defined class and if this change favors forming a tighter entrained class (i.e., flipping effect). We can evaluate the phase synchronization changes across weeks by comparing the entrainment and flipping effects. Circular clustering and two introduced effects are evaluated in Rstudio (package 'OptCirClust' [21]: https://cran.r-project.org/web/packages/OptCirClust/index.html and package 'circular' [22]: https://cran.r-project.org/web/packages/circular/index.html).

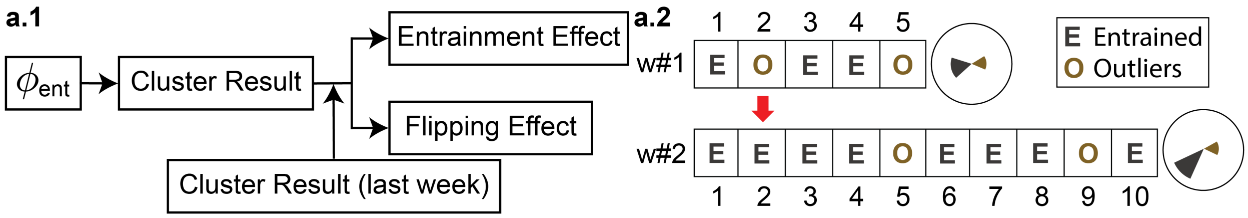


**Fig. S.5.** Details of the phase synchronization measurement. **a.1** Flowchart of the phase synchronization measurement by treatment week. Entrainment and flipping effects were measured by comparing the clustering result of $\phi_{ent}$ until the current week with the previous week. **a.2** Example of entrainment and flipping effect calculation. Entrainment and flipping effect are determined by the number of sessions in each class (entrained vs. outlier) and tightness of the formed class (i.e., the standard deviation of $\phi_{ent}$ within each defined class).

### S.12. CAUSALITY BETWEEN EXCITABILITY AND ENTRAINMENT

Based on the result of the Granger causality test [23], we did not find excitability/entrainment helps forecast entrainment/excitability on a group-level analysis. However, this conclusion might be limited by the small number of subjects and observations (one averaged value each week and six weeks in total), so further experimentation is needed to determine if there is a causal relationship between excitability and entrainment. Nevertheless, for the prediction of treatment outcomes, according to the result of this study, we think entrainment plays a more critical role than excitability because when we use the predictive diagram introduced in Figure 5 of the main text without the excitability threshold, 5 of 6 subjects within the SYNC responder group will still be selected to finish the entire treatment (response rate: 83.3%).
